# Novel risk loci for COVID-19 hospitalization among admixed American populations

**DOI:** 10.1101/2023.08.11.23293871

**Authors:** Silvia Diz-de Almeida, Raquel Cruz, Andre D. Luchessi, José M. Lorenzo-Salazar, Miguel López de Heredia, Inés Quintela, Rafaela González-Montelongo, Vivian N. Silbiger, Marta Sevilla Porras, Jair Antonio Tenorio Castaño, Julian Nevado, Jose María Aguado, Carlos Aguilar, Sergio Aguilera-Albesa, Virginia Almadana, Berta Almoguera, Nuria Alvarez, Álvaro Andreu-Bernabeu, Eunate Arana-Arri, Celso Arango, María J. Arranz, Maria-Jesus Artiga, Raúl C. Baptista-Rosas, María Barreda- Sánchez, Moncef Belhassen-Garcia, Joao F. Bezerra, Marcos A.C. Bezerra, Lucía Boix-Palop, María Brion, Ramón Brugada, Matilde Bustos, Enrique J. Calderón, Cristina Carbonell, Luis Castano, Jose E. Castelao, Rosa Conde-Vicente, M. Lourdes Cordero-Lorenzana, Jose L. Cortes-Sanchez, Marta Corton, M. Teresa Darnaude, Alba De Martino-Rodríguez, Victor del Campo-Pérez, Aranzazu Diaz de Bustamante, Elena Domínguez-Garrido, Rocío Eirós, María Carmen Fariñas, María J. Fernandez-Nestosa, Uxía Fernández-Robelo, Amanda Fernández-Rodríguez, Tania Fernández-Villa, Manuela Gago-Domínguez, Belén Gil-Fournier, Javier Gómez-Arrue, Beatriz González Álvarez, Fernan Gonzalez Bernaldo de Quirós, Anna González-Neira, Javier González-Peñas, Juan F. Gutiérrez-Bautista, María José Herrero, Antonio Herrero-Gonzalez, María A. Jimenez-Sousa, María Claudia Lattig, Anabel Liger Borja, Rosario Lopez-Rodriguez, Esther Mancebo, Caridad Martín-López, Vicente Martín, Oscar Martinez-Nieto, Iciar Martinez-Lopez, Michel F. Martinez-Resendez, Ángel Martinez-Perez, Juliana F. Mazzeu, Eleuterio Merayo Macías, Pablo Minguez, Victor Moreno Cuerda, Silviene F. Oliveira, Eva Ortega-Paino, Mara Parellada, Estela Paz-Artal, Ney P.C. Santos, Patricia Pérez-Matute, Patricia Perez, M. Elena Pérez-Tomás, Teresa Perucho, Mel·lina Pinsach-Abuin, Guillermo Pita, Ericka N. Pompa-Mera, Gloria L. Porras-Hurtado, Aurora Pujol, Soraya Ramiro León, Salvador Resino, Marianne R. Fernandes, Emilio Rodríguez-Ruiz, Fernando Rodriguez-Artalejo, José A. Rodriguez-Garcia, Francisco Ruiz-Cabello, Javier Ruiz-Hornillos, Pablo Ryan, José Manuel Soria, Juan Carlos Souto, Eduardo Tamayo, Alvaro Tamayo-Velasco, Juan Carlos Taracido-Fernandez, Alejandro Teper, Lilian Torres-Tobar, Miguel Urioste, Juan Valencia-Ramos, Zuleima Yáñez, Ruth Zarate, Itziar de Rojas, Agustín Ruiz, Pascual Sánchez, Luis Miguel Real, SCOURGE Cohort Group, Encarna Guillen-Navarro, Carmen Ayuso, Esteban Parra, José A. Riancho, Augusto Rojas-Martinez, Carlos Flores, Pablo Lapunzina, Ángel Carracedo

**Author notes:** These authors contributed equally: Carlos Flores, Pablo Lapunzina, Ángel Carracedo. These authors contributed equally: Silvia Diz-de Almeida, Raquel Cruz.

## Abstract

The genetic basis of severe COVID-19 has been thoroughly studied, and many genetic risk factors shared between populations have been identified. However, reduced sample sizes from non-European groups have limited the discovery of population-specific common risk loci. In this second study nested in the SCOURGE consortium, we conducted a GWAS for COVID-19 hospitalization in admixed Americans, comprising a total of 4,702 hospitalized cases recruited by SCOURGE and seven other participating studies in the COVID-19 Host Genetic Initiative. We identified four genome-wide significant associations, two of which constitute novel loci and were first discovered in Latin American populations (*BAZ2B* and *DDIAS*). A trans-ethnic meta-analysis revealed another novel cross-population risk locus in *CREBBP*. Finally, we assessed the performance of a cross-ancestry polygenic risk score in the SCOURGE admixed American cohort. This study constitutes the largest GWAS for COVID-19 hospitalization in admixed Latin Americans conducted to date. This allowed to reveal novel risk loci and emphasize the need of considering the diversity of populations in genomic research.

## Introduction

To date, more than 50 loci associated with COVID-19 susceptibility, hospitalization, and severity have been identified using genome-wide association studies (GWAS)^1,2^. The COVID-19 Host Genetics Initiative (HGI) has made significant efforts^3^ to augment the power to identify disease loci by recruiting individuals from diverse populations and conducting a trans-ancestry meta-analysis. Despite this, the lack of genetic diversity and a focus on cases of European ancestries still predominate in the studies^4,5^. In addition, while trans-ancestry meta-analyses are a powerful approach for discovering shared genetic risk variants with similar effects across populations^6^, they may fail to identify risk variants that have larger effects on particular underrepresented populations. Genetic disease risk has been shaped by the particular evolutionary history of populations and environmental exposures^7^. Their action is particularly important for infectious diseases due to the selective constraints that are imposed by host □ pathogen interactions^8,9^. Literature examples of this in COVID-19 severity include a *DOCK2* gene variant in East Asians^10^ and frequent loss-of-function variants in *IFNAR1* and *IFNAR2* genes in Polynesian and Inuit populations, respectively^11,12^.

Including diverse populations in case□control GWAS studies with unrelated participants usually requires a prior classification of individuals in genetically homogeneous groups, which are typically analyzed separately to control the population stratification effects^13^. Populations with recent admixture impose an additional challenge to GWASs due to their complex genetic diversity and linkage disequilibrium (LD) patterns, requiring the development of alternative approaches and a careful inspection of results to reduce false positives due to population structure^7^. In fact, there are benefits in study power from modeling the admixed ancestries either locally, at the regional scale in the chromosomes, or globally, across the genome, depending on factors such as the heterogeneity of the risk variant in frequencies or the effects among the ancestry strata^14^. Despite the development of novel methods specifically tailored for the analysis of admixed populations^15^, the lack of a standardized analysis framework and the difficulties in confidently clustering admixed individuals into particular genetic groups often leads to their exclusion from GWAS.

The Spanish Coalition to Unlock Research on Host Genetics on COVID-19 (SCOURGE) recruited COVID-19 patients between March and December 2020 from hospitals across Spain and from March 2020 to July 2021 in Latin America (https://www.scourge-covid.org). A first GWAS of COVID-19 severity among Spanish patients of European descent revealed novel disease loci and explored age- and sex-varying effects of genetic factors^16^. Here, we present the findings of a GWAS meta-analysis in admixed Latin American (AMR) populations, comprising individuals from the SCOURGE Latin-American cohort and the HGI studies, which allowed us to identify two novel severe COVID-19 loci, *BAZ2B* and *DDIAS*. Further analyses modeling the admixture from three genetic ancestral components and performing a trans-ethnic meta-analysis led to the identification of an additional risk locus near *CREBBP*. We finally assessed a cross-ancestry polygenic risk score model with variants associated with critical COVID-19.

## Results

### Meta-analysis of COVID-19 hospitalization in admixed Americans

#### Study cohorts

Within the SCOURGE consortium, we included 1,608 hospitalized cases and 1,887 controls (not hospitalized COVID-19 patients) from Latin American countries and from recruitments of individuals of Latin American descent conducted in Spain (Supplementary Table 1). Quality control details and estimation of global genetic inferred ancestry (GIA) (Supplementary Figure 1) are described in Methods, whereas clinical and demographic characteristics of patients included in the analysis are shown in Table 1. Summary statistics from the SCOURGE cohort were obtained under a logistic mixed model with the SAIGE model (Methods). Another seven studies participating in the COVID-19 HGI consortium were included in the meta-analysis of COVID-19 hospitalization in admixed Americans (Figure 1).

**Table 1.** Demographic characteristics of the SCOURGE Latin American cohort.

| Variable | Non Hospitalized<br>N = 1,887 | Hospitalized<br>N = 1,625 |
| --- | --- | --- |
| Age – mean years ± SD | 39.1 ± 11.9 | 54.1 ± 14.5 |
| Sex - N (%) |  |  |
| Female (%) | 1253 (66.4) | 668 (41.1) |
| GIA* – % mean ±SD | | |
| European | 54.4 ± 16.2 | 39.4 ± 20.7 |
| African | 15.3 ± 12.7 | 9.1 ± 11.6 |
| Native American | 30.3 ± 19.8 | 51.3 ± 26.5 |
| Comorbidities - N (%) |  |  |
| Vascular/endocrinological | 488 (25.9) | 888 (64.5) |
| Cardiac | 60 (3.2) | 151 (9.3) |
| Nervous | 15 (0.8) | 61 (3.8) |
| Digestive | 14 (0.7) | 33 (2.0) |
| Onco-hematological | 21 (1.1) | 48 (3.00) |
| Respiratory | 76 (4.0) | 118 (7.3) |

\*Global genetic inferred ancestry.

**Figure 1.**
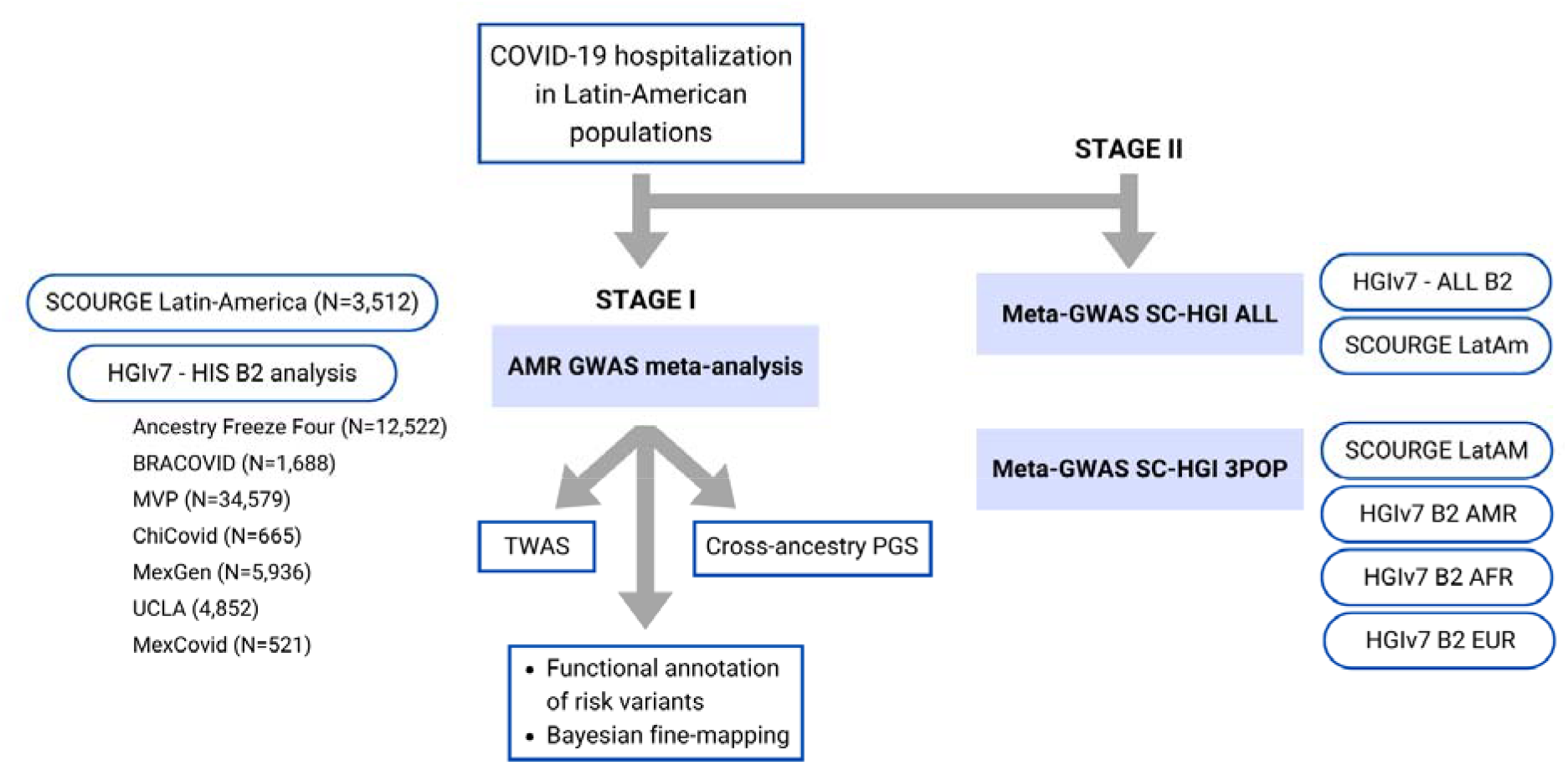
Flow chart of this study.

#### GWAS meta-analysis

We performed a fixed-effects GWAS meta-analysis using the inverse of the variance as weights for the overlapping markers. The combined GWAS sample size consisted of 4,702 admixed AMR hospitalized cases and 68,573 controls.

This GWAS meta-analysis revealed genome-wide significant associations at four risk loci (Table 2, Figure 2), two of which (*BAZ2B* and *DDIAS*) were novel discoveries. Four lead variants were identified, linked to other 310 variants (Supplementary Tables 2-3). A gene-based association test revealed a significant association in *BAZ2B* and in previously known COVID-19 risk loci: *LZTFL1, XCR1, FYCO1, CCR9*, and *IFNAR2* (Supplementary Table 4).

**Table 2.**
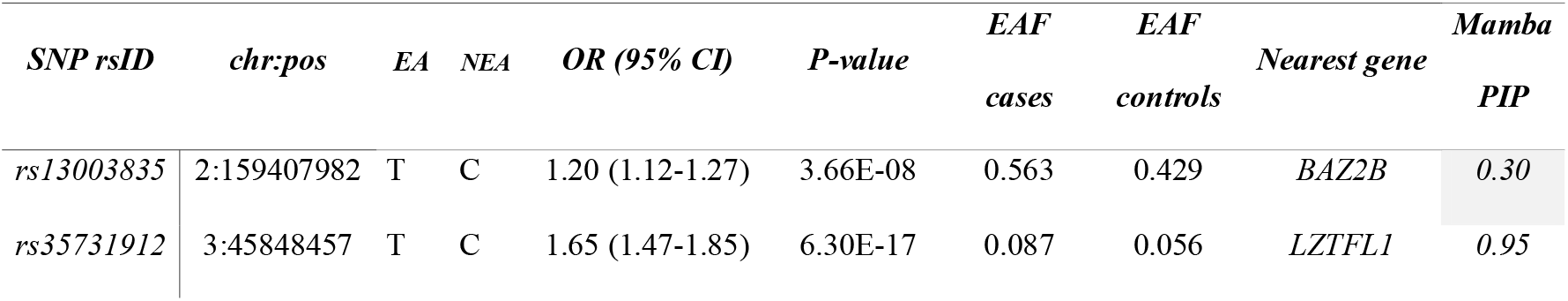
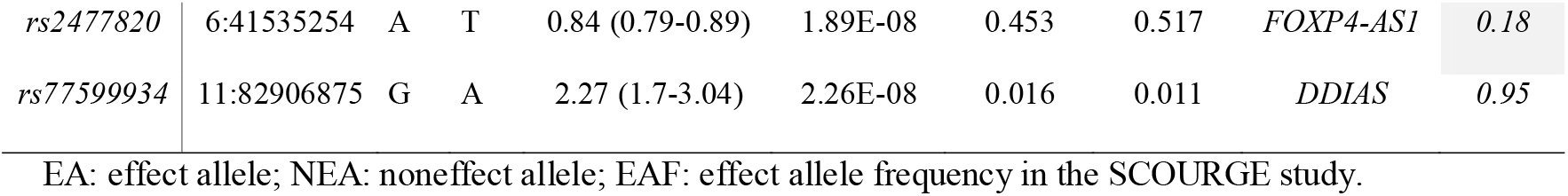
Lead independent variants in the admixed AMR GWAS meta-analysis.

**Table 3.**
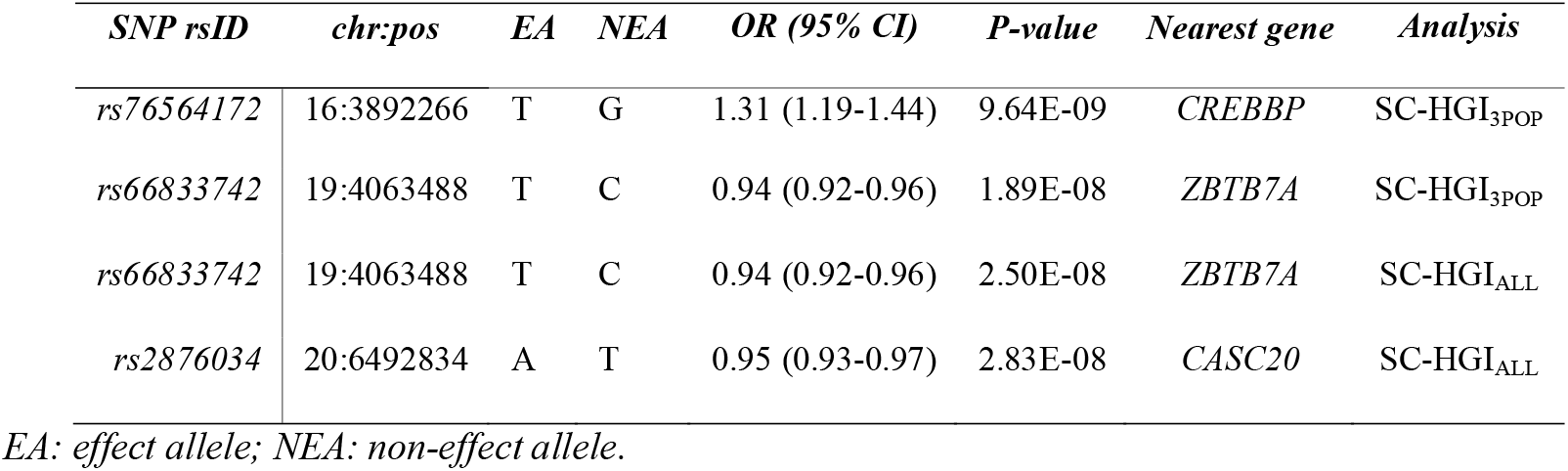
Novel variants in the SC-HGI_ALL_ and SC-HGI_3POP_ meta-analyses (with respect to HGIv7). Independent signals after LD clumping.

| *SNP rsID* | *chr:pos* | *EA* | *NEA* | *OR (95% CI)* | *P-value* | *Nearest gene* | *Analysis* |
| --- | --- | --- | --- | --- | --- | --- | --- |
| *rs76564172* | 16:3892266 | T | G | 1.31 (1.19-1.44) | 9.64E-09 | *CREBBP* | SC-HGI<sub>3POP</sub> |
| *rs66833742* | 19:4063488 | T | C | 0.94 (0.92-0.96) | 1.89E-08 | *ZBTB7A* | SC-HGI<sub>3POP</sub> |
| *rs66833742* | 19:4063488 | T | C | 0.94 (0.92-0.96) | 2.50E-08 | *ZBTB7A* | SC-HGI<sub>ALL</sub> |
| *rs2876034* | 20:6492834 | A | T | 0.95 (0.93-0.97) | 2.83E-08 | *CASC20* | SC-HGI<sub>ALL</sub> |

EA: effect allele; NEA: non-effect allele.

**Figure 2.**
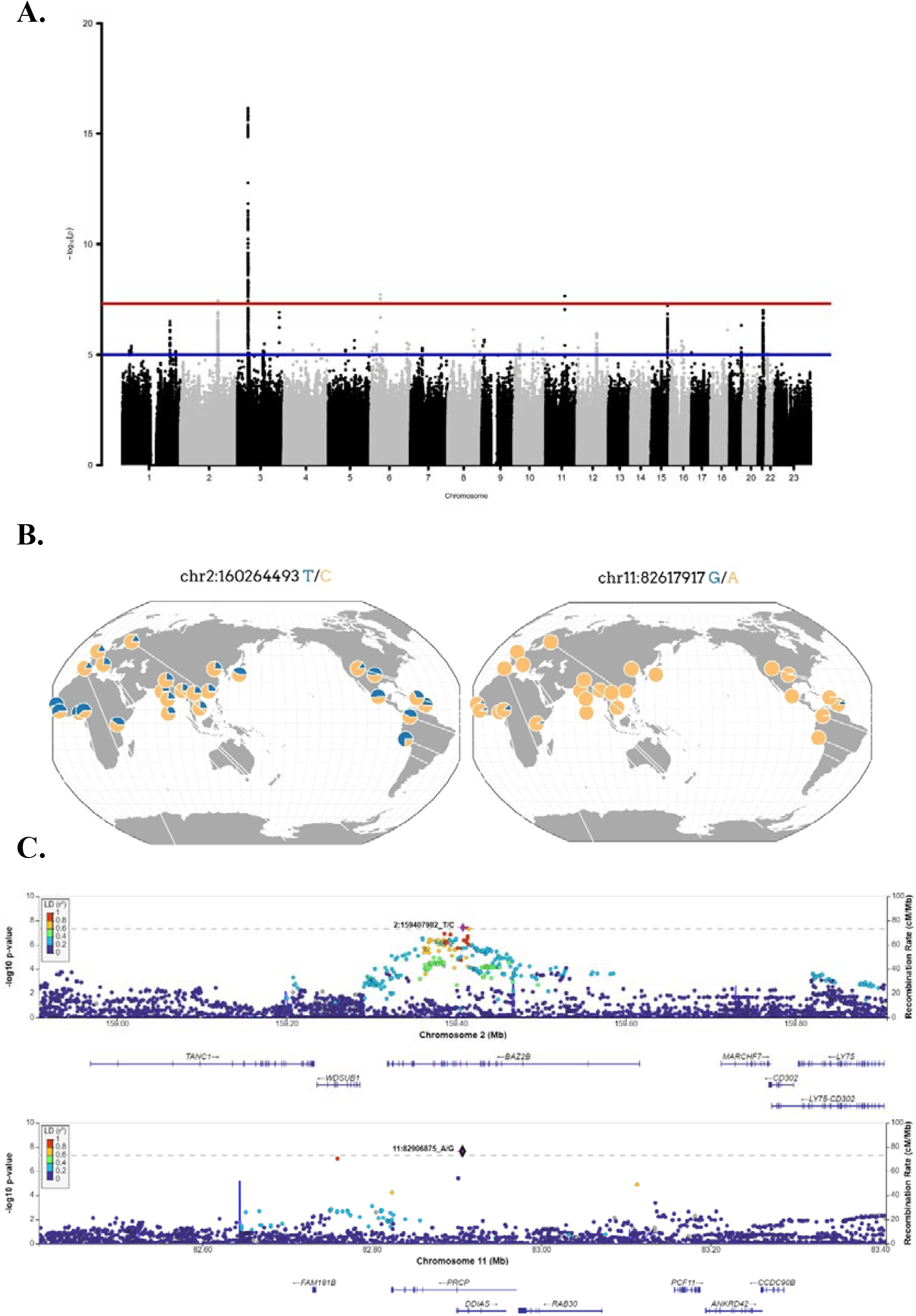
**A) Manhattan plot for the admixed AMR GWAS meta-analysis.** Probability thresholds at p=5×10^−8^ and p=5×10^−5^ are indicated by the horizontal lines. Genome-wide significant associations with COVID-19 hospitalizations were found on chromosome 2 (within *BAZ2B*), chromosome 3 (within *LZTFL1*), chromosome 6 (within *FOXP4*), and chromosome 11 (within *DDIAS*). A Quantile □ Quantile plot is shown in supplementary Figure 2. **B) Regional association plots for rs1003835 at chromosome 2 and rs77599934 at chromosome 11**; **C) Allele frequency distribution across the 1000 Genomes Project populations for the lead variants rs1003835 and rs77599934**. Retrieved from *The Geography of Genetic Variants Web* or GGV.

Located within the *BAZ2B* gene, the sentinel variant rs13003835 is an intronic variant associated with an increased risk of COVID-19 hospitalization (Odds Ratio [OR]=1.20, 95% Confidence Interval [CI]=1.12-1.27, p=3.66×10^−8^). This association was not previously reported in any GWAS of COVID-19 published to date. Interestingly, rs13003835 did not reach significance (p=0.972) in the COVID-19 HGI trans-ancestry meta-analysis including the five population groups^1^.

The other novel risk locus is led by the sentinel variant rs77599934, a rare intronic variant located in chromosome 11 within *DDIAS* and associated with the risk of COVID-19 hospitalization (OR=2.27, 95% CI=1.70-3.04, p=2.26×10^−8^).

We also observed a suggestive association with rs2601183 in chromosome 15, which is located between *ZNF774* and *IQGAP1* (allele-G OR=1.20, 95% CI=1.12-1.29, p=6.11×10^−8^, see Supplementary Table 2), which has not yet been reported in any other GWAS of COVID-19 to date.

The GWAS meta-analysis also pinpointed two significant variants at known loci, *LZTFL1* and *FOXP4*. The SNP rs35731912 was previously associated with COVID-19 severity in EUR populations^17^, and it was mapped to *LZTFL1*. While rs2477820 is a novel risk variant within the *FOXP4 gene*, it has a moderate LD (*r*^2^=0.295) with rs2496644, which has been linked to COVID-19 hospitalization^18^. This is consistent with the effects of LD in tag-SNPs when conducting GWAS in diverse populations.

None of the lead variants was associated with the comorbidities included in Table 1.

### Functional mapping of novel risk variants

Variants belonging to the lead loci were prioritized by positional and expression quantitative trait loci (eQTL) mapping with FUMA, resulting in 31 mapped genes (Supplementary Table 5). Within the region surrounding the lead variant in chromosome 2, FUMA prioritized four genes in addition to *BAZ2B* (*PLA2R1, LY75, WDSUB1*, and *CD302*). rs13003835 (allele C) is an eQTL of *LY75* in the esophagus mucosa (NES=0.27) and of *BAZ2B-AS* in whole blood (NES=0.27), while rs2884110 (R^2^=0.85) is an eQTL of *LY75* in lung (NES=0.22). As for the chromosome 11, rs77599934 (allele G) is in moderate-to-strong LD (r^2^=0.776) with rs60606421 (G deletion, allele -) which is an eQTL associated with a reduced expression of *DDIAS* in the lungs (NES=-0.49, allele -). Associations with expression are shown in the supplementary Figure 4. The sentinel variant for the region in chromosome 16 is in perfect LD (*r*^2^=1) with rs601183, an eQTL of *ZNF774* in the lung.

#### Bayesian fine mapping

We performed different approaches to narrow down the prioritized loci to a set of most likely genes driving the associations. First, we computed credible sets at the 95% confidence level for causal variants and annotated them with VEP (and V2G aggregate scoring. The 95% confidence credible set from the region of chromosome 2 around rs13003835 included 76 variants, which can be found in the Supplementary Table 6 (VEP and V2G annotations are included in the Supplementary Tables 7 and 8). TheV2G score prioritized *BAZ2B* as the most likely gene driving the association. However, the approach was unable to converge allocating variants in a 95% confidence credible set for the region in chromosome 11.

#### Transcriptome-wide association study (TWAS)

Five novel genes, namely, *SLC25A37, SMARCC1, CAMP, TYW3*, and *S100A12* (supplementary Table 9), were found to be significantly associated in the cross-tissue TWAS. To our knowledge, these genes have not been reported previously in any COVID-19 TWAS or GWAS analyses published to date. In the single tissue analyses, *ATP5O* and *CXCR6* were significantly associated in the lungs, *CCR9* was significantly associated in whole blood, and *IFNAR2* and *SLC25A37* were associated in lymphocytes.

Likewise, we carried out TWAS analyses using the models trained in the admixed populations. However, no significant gene pairs were detected in this case. The top 10 genes with the lowest p-values for each of the datasets (Puerto-Ricans, Mexicans, African-Americans and pooled cohorts) are shown in the Supplementary Table 10. Although not significant, *KCNC3* was repeated in the four analyses, whereas *MAPKAPK3, NAPSA* and *THAP5* were repeated in 3 out of 4. Both *NAPSA* and *KCNC3* are located in the chromosome 19 and were reported in the latest HGI meta-analysis^19^.

All mapped genes from analyses conducted in AMR populations are shown in Figure 5.

### Genetic architecture of COVID-19 hospitalization in AMR populations

#### Allele frequencies of rs13003835 and rs77599934 across ancestries

Neither rs13003835 (*BAZ2B*) nor rs77599934 (*DDIAS*) were significantly associated in the COVID-19 HGI B2 cross-population or population-specific meta-analyses. Thus, we investigated their allele frequencies (AF) across populations and compared their effect sizes.

According to gnomAD v3.1.2, the T allele at rs13003835 (*BAZ2B*) has an AF of 43% in admixed AMR groups, while AF is lower in the EUR populations (16%) and in the global sample (29%). Local ancestry inference (LAI) reported by gnomAD shows that within the Native-American component, the risk allele T is the major allele, whereas it is the minor allele within the African and European LAI components. These large differences in AF might be the reason underlying the association found in AMR populations. However, when comparing effect sizes between populations, we found that they were in opposite directions between SAS-AMR and EUR-AFR-EAS and that there was large heterogeneity among them (Figure 3). We queried SNPs within 50kb windows of the lead variant in each of the other populations that had p-value<0.01. The variant with the lowest p-value in the EUR population was rs559179177 (p=1.72×10^−4^), which is in perfect LD (r^2^=1) in the 1KGP EUR population with our sentinel variant (rs13003835), and in moderate LD r^2^=0.4 in AMR populations. Since this variant was absent from the AMR analysis, probably due to its low frequency, it could not be meta-analyzed. Power calculations revealed that the EUR analysis was underpowered for this variant to achieve genome-wide significance (77.6%, assuming an effect size of 0.46, EAF= 0.0027, and number of cases/controls as shown in the HGI website for B2-EUR). In the cross-population meta-analysis (B2-ALL), rs559179177 obtained a p-value of 5.9×10^−4^.

**Figure 3.**
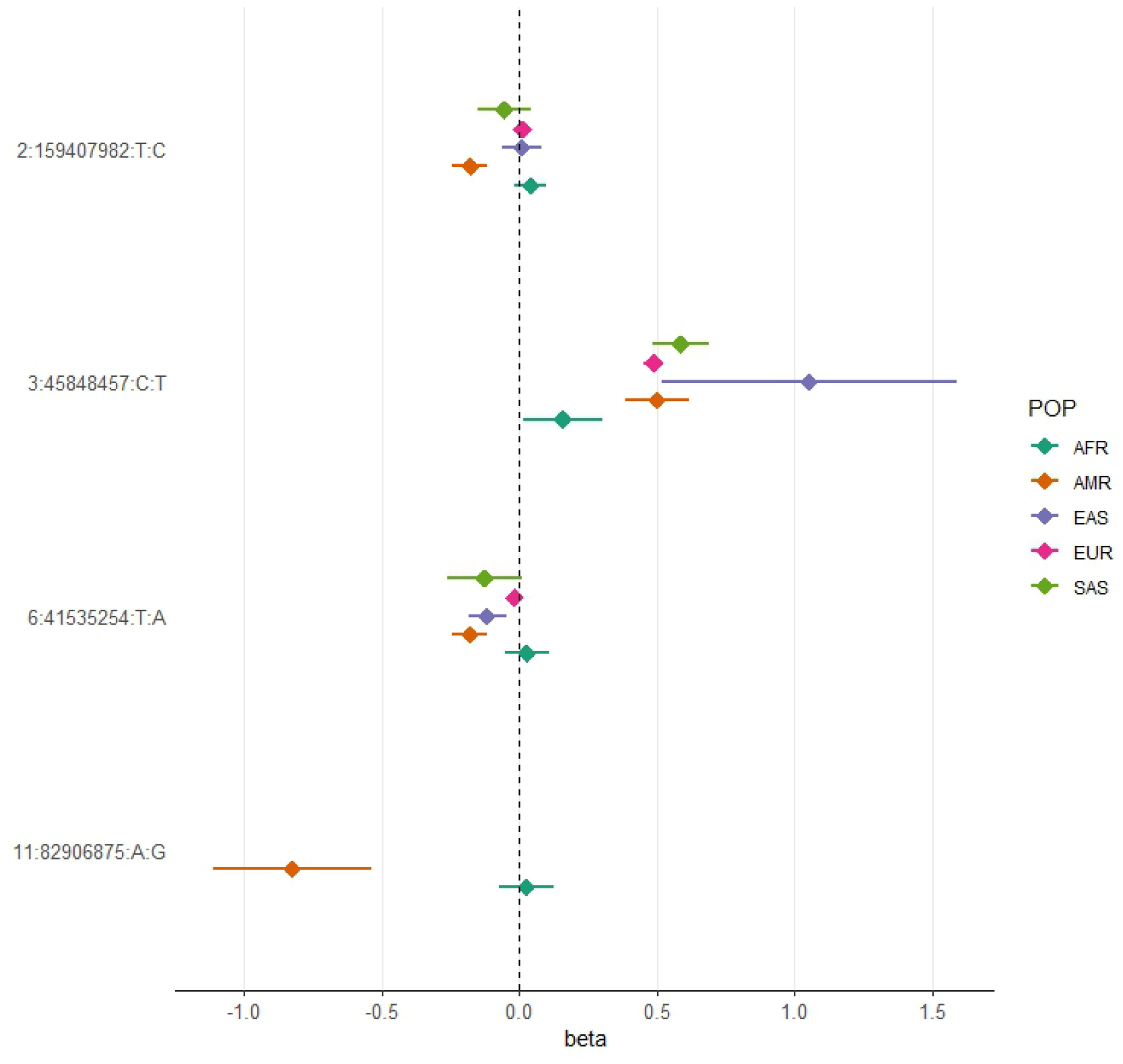
Forest plot showing effect sizes and the corresponding confidence intervals for the sentinel variants identified in the AMR meta-analysis across populations. All beta values with their corresponding CIs were retrieved from the B2 population-specific meta-analysis from the HGI v7 release, except for AMR, for which the beta value and IC from the HGI_AMR_-SCOURGE meta-analysis are represented.

rs77599934 (*DDIAS*) had an AF of 1.1% for the G allele in the nonhospitalized controls (Table 2), in line with the recorded gnomAD AF of 1% in admixed AMR groups. This variant has the potential to be a population-specific variant, given the allele frequencies in other population groups, such as EUR (0% in Finnish, 0.025% in non-Finnish), EAS (0%) and SAS (0.042%), and its greater effect size over AFR populations (Figure 3). Examining the LAI, the G allele occurs at a 10.8% frequency in the African component, while it is almost absent in the Native-American and European. Due to its low MAF, rs77599934 was not analyzed in the COVID-19 HGI B2 cross-population meta-analysis and was only present in the HGI B2 AFR population-specific meta-analysis, precluding the comparison (Figure 3). For this reason, we retrieved the variant with the lowest p-value within a 50 kb region around rs77599934 in the COVID-19 HGI cross-population analysis to investigate whether it was in moderate-to-strong LD with our sentinel variant. The variant with the smallest p-value was rs75684040 (OR=1.07, 95% CI=1.03-1.12, p=1.84×10^−3^). However, LD calculations using the 1KGP phase 3 dataset indicated that rs77599934 and rs75684040 were poorly correlated (r^2^=0.11). As for AFR populations, the variant with the lowest p-value was rs138860115 (p=8.3×10^−3^), but it was not correlated with the lead SNP of this locus.

#### Cross-population meta-analyses

We carried out two cross-ancestry inverse variance-weighted fixed-effects meta-analyses with the admixed AMR GWAS meta-analysis results to evaluate whether the discovered risk loci replicated when considering other population groups. In doing so, we also identified novel cross-population COVID-19 hospitalization risk loci.

First, we combined the SCOURGE Latin American GWAS results with the HGI B2 ALL analysis (supplementary Table 11). We refer to this analysis as the SC-HGI_ALL_ meta-analysis. Out of the 40 genome-wide significant loci associated with COVID-19 hospitalization in the last HGI release^1^, this study replicated 39, and the association was stronger than in the original study in 29 of those (supplementary Table 12). However, the variant rs13003835 located in *BAZ2B* did not replicate (OR=1.00, 95% CI=0.98-1.03, p=0.644).

In this cross-ancestry meta-analysis, we replicated two associations that were not found in HGIv7, albeit they were sentinel variants in the latest GenOMICC meta-analysis^2^. We found an association at the *CASC20* locus led by the variant rs2876034 (OR=0.95, 95% CI=0.93-0.97, p=2.83×10^−8^). This variant is in strong LD with the sentinel variant of that study (rs2326788, *r*^2^=0.92), which was associated with critical COVID-19^2^. In addition, this meta-analysis identified the variant rs66833742 near *ZBTB7A* associated with COVID-19 hospitalization (OR=0.94, 95% CI=0.92-0.96, p=2.50×10^−8^). Notably, rs66833742 or its perfect proxy rs67602344 (r^2^=1) are also associated with upregulation of *ZBTB7A* in whole blood and in esophageal mucosa. This variant was previously associated with COVID-19 hospitalization^2^.

In a second analysis, we also explored the associations across the defined admixed AMR, EUR, and AFR ancestral sources by combining through meta-analysis the SCOURGE Latin American GWAS results with the HGI studies in EUR, AFR, and admixed AMR and excluding those from EAS and SAS (supplementary Table 13). We refer to this as the SC-HGI_3POP_ meta-analysis. The association at rs13003835 (*BAZ2B*, OR=1.01, 95% CI=0.98-1.03, p=0.605) was not replicated, and rs77599934 near *DDIAS* could not be assessed, although the association at the *ZBTB7A* locus was confirmed (rs66833742, OR=0.94, 95% CI=0.92-0.96, p=1.89×10^−8^). The variant rs76564172 located near *CREBBP* also reached statistical significance (OR=1.31, 95% CI=1.25-1.38, p=9.64×10^−9^). The sentinel variant of the region linked to *CREBBP* (in the trans-ancestry meta-analysis) was also subjected a Bayesian fine mapping (supplementary Table 6). Eight variants were included in the credible set for the region in chromosome 16 (meta-analysis SC-HGI_3POP_).

### Polygenic risk score models

Using the 49 variants associated with disease severity that are shared across populations according to the HGIv7, we constructed a polygenic risk score (PGS) model to assess its generalizability in the admixed AMR (supplementary Table 14). First, we calculated the PGS for the SCOURGE Latin Americans and explored the association with COVID-19 hospitalization under a logistic regression model. The PGS model was associated with a 1.48-fold increase in COVID-19 hospitalization risk per every PGS standard deviation. It also contributed to explaining a slightly larger variance (ΔR2=1.07%) than the baseline model. Subsequently, we divided the individuals into PGS deciles and percentiles to assess their risk stratification. The median percentile among controls was 40, while in cases, it was 63. Those in the top PGS decile exhibited a 2.89-fold (95% CI=2.37-3.54, p=1.29×10^−7^) greater risk compared to individuals in the deciles between 4 and 6 (corresponding to a score of the median distribution).

We also examined the distribution of PGS scores across a 5-level severity scale to further determine if there was any correspondence between clinical severity and genetic risk. Median PGS scores were lower in the asymptomatic and mild groups, whereas higher median scores were observed in the moderate, severe, and critical patients (Figure 4). We fitted a multinomial model using the asymptomatic class as a reference and calculated the OR for each category (supplementary Table 15), observing that the disease genetic risk was similar among asymptomatic, mild, and moderate patients. Given that the PGS was built with variants associated with critical disease and/or hospitalization and that the categories severe and critical correspond to hospitalized patients, these results underscore the ability of cross-ancestry PGS for risk stratification even in an admixed population.

**Figure 4.**
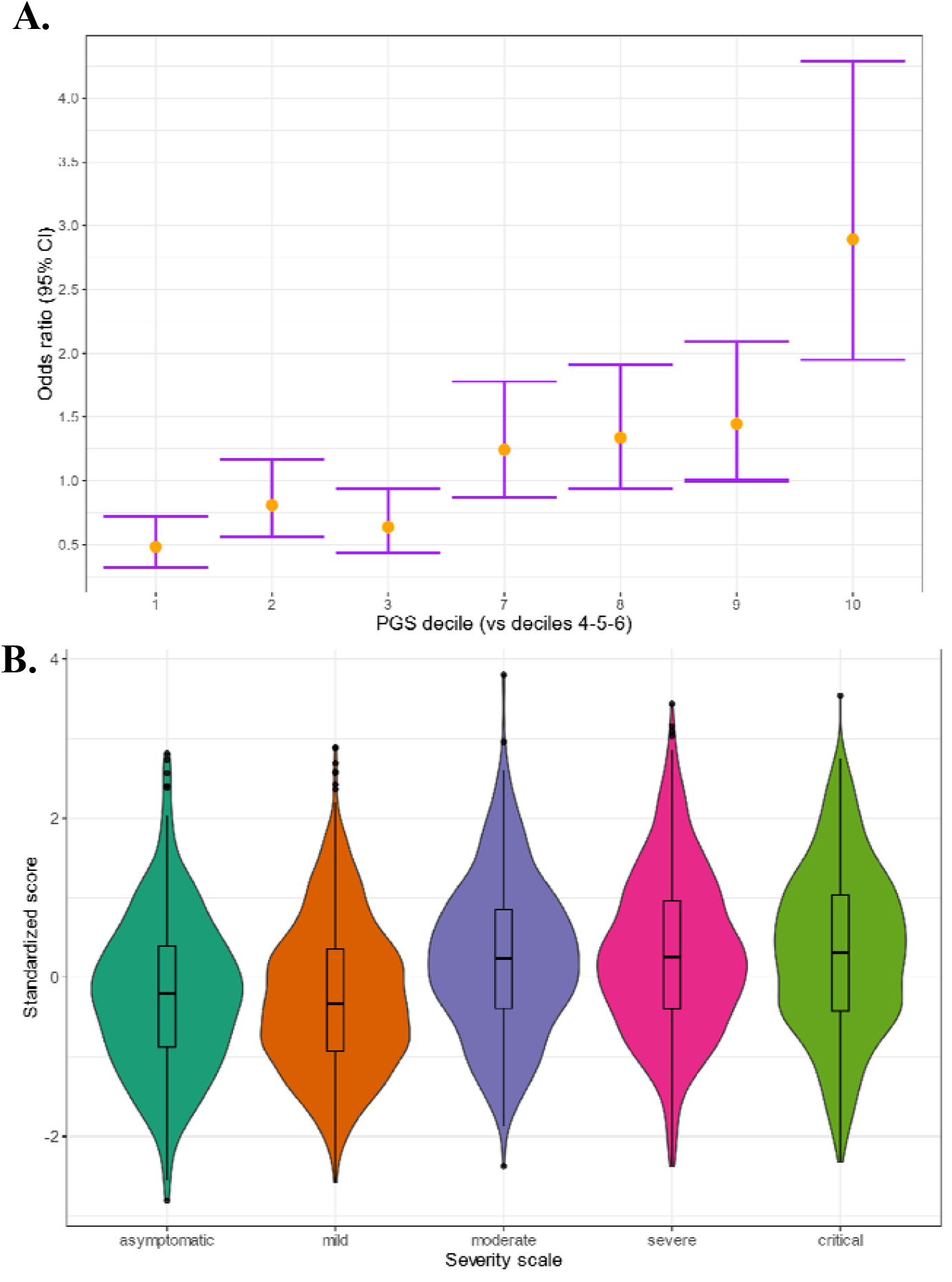
(A) Polygenic risk stratified by PGS deciles comparing each risk group against the lowest risk group (OR-95% CI); (B) Distribution of the PGS scores in each of the severity scale classes. 0-Asymptomatic, 1-Mild disease, 2-Moderate disease, 3-Severe disease, 4-Critical disease.

**Figure 5.**
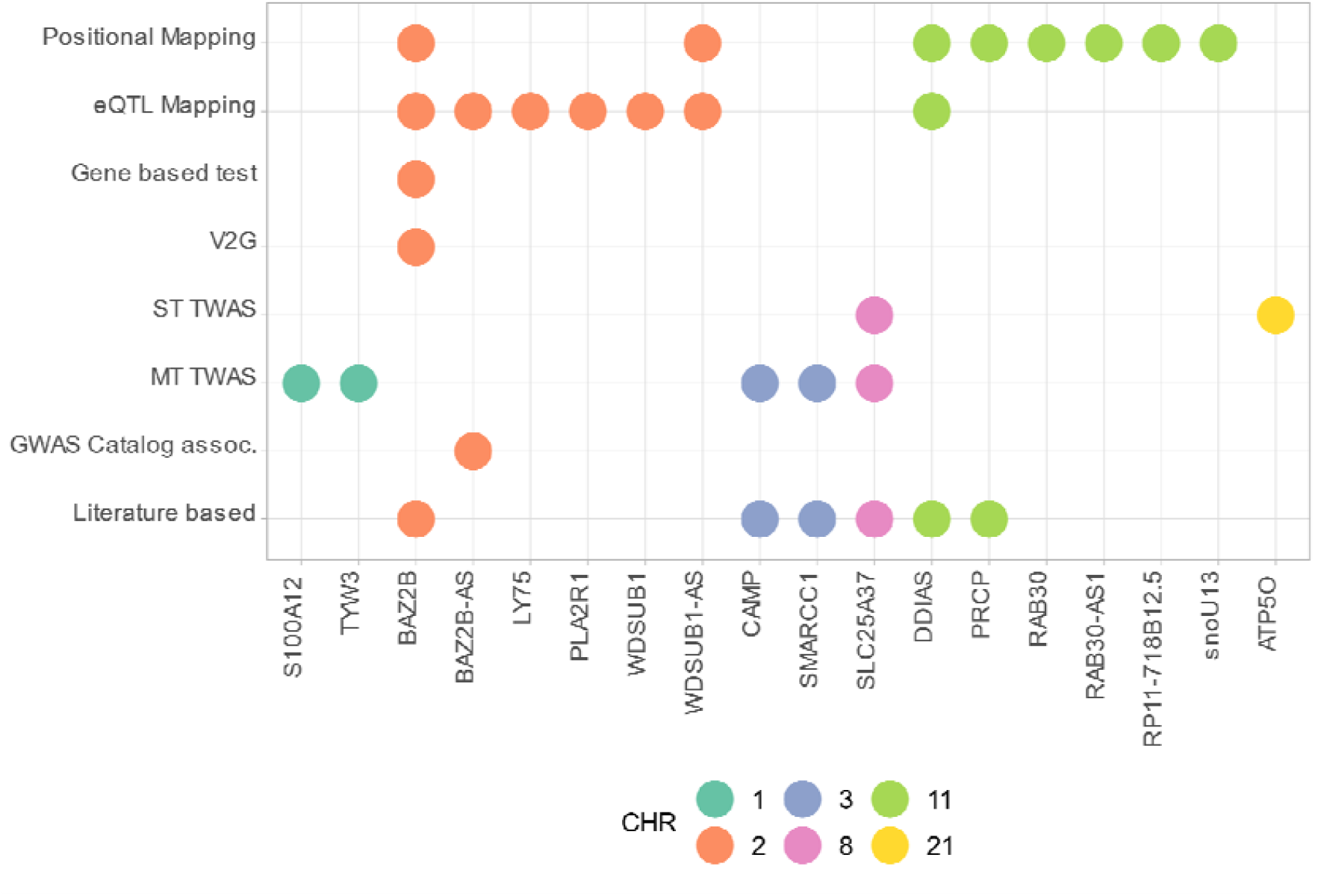
Summary of the results from gene prioritization strategies used for genetic associations in AMR populations. GWAS catalog association for *BAZ2B-AS* was with FEV/FCV ratio. Literature based evidence is further explored in discussion.

## DISCUSSION

We have conducted the largest GWAS meta-analysis of COVID-19 hospitalization in admixed AMR to date. While the genetic risk basis discovered for COVID-19 is largely shared among populations, trans-ancestry meta-analyses on this disease have primarily included EUR samples. This dominance of GWAS in Europeans and the subsequent bias in sample sizes can mask population-specific genetic risks (i.e., variants that are monomorphic in some populations) or be less powered to detect risk variants having higher allele frequencies in population groups other than Europeans. In this sense, after combining data from admixed AMR patients, we found two risk loci that were first discovered in a GWAS of Latin American populations. Interestingly, the sentinel variant rs77599934 in the *DDIAS* gene is a rare coding variant (∼1% for allele G) with a large effect on COVID-19 hospitalization that is nearly monomorphic in most of the other populations. This has likely led to its exclusion from the cross-population meta-analyses conducted to date, remaining undetectable.

Fine mapping of the region harboring *DDIAS* did not reveal further information about which gene could be the more prone to be causal or about the functional consequences of the risk variant, but our sentinel variant was in strong LD with an eQTL that associated with reduced gene expression of *DDIAS* in the lung. *DDIAS*, known as damage-induced apoptosis suppressor gene, is itself a plausible candidate gene. It has been linked to DNA damage repair mechanisms: research has shown that depletion of *DDIAS* leads to an increase in ATM phosphorylation and the formation of p53-binding protein (53BP1) foci, a known biomarker of DNA double-strand breaks, suggesting a potential role in double-strand break repair^20^. Interestingly, a study found that infection by SARS-CoV-2 also triggered the phosphorylation of ATM kinase and inhibited repair mechanisms, causing the accumulation of DNA damage^21^. This gene has also been proposed as a potential biomarker for lung cancer after finding that it interacts with STAT3 in lung cancer cells, regulating IL-6^22,23^ and thus mediating inflammatory processes, while another study determined that its blockade inhibited lung cancer cell growth^24^.Another prioritized gene from this region was *PRCP*, an angiotensinase which shares substrate specificity with ACE2 receptor. It has been positively linked to hypertension and some studies have raised hypotheses on its role in COVID-19 progression, particularly in relation to the development of pro-thrombotic events ^25,26^.

The risk region found in chromosome 2 harbors more than one gene. The lead variant rs13003835 is located within *BAZ2B*, and it increases the expression of the antisense *BAZ2B* gene in whole blood. *BAZ2B* encodes one of the regulatory subunits of the Imitation switch (ISWI) chromatin remodelers^27^ constituting the BRF-1/BRF-5 complexes with SMARCA1 and SMARCA5, respectively. Interestingly, it was discovered that *lnc-BAZ2B* promotes macrophage activation through regulation of *BAZ2B* expression. Its over-expression resulted in pulmonary inflammation and elevated levels of *MUC5AC* in mice with asthma^28^. This variant was also an eQTL for *LY75* (encoding lymphocyte antigen 75) in the esophageal mucosa tissue. Lymphocyte antigen 75 is involved in immune processes through antigen presentation in dendritic cells and endocytosis^29^ and has been associated with inflammatory diseases, representing a compelling candidate for the region. Increased expression of *LY75* has been detected within hours after infection by SARS-CoV-2^30,31^. It is worth noting that differences in AF for this variant suggest that analyses in AMR populations might be more powered to detect the association, supporting the necessity of population-specific studies.

A third novel risk region was observed on chromosome 15 between the *IQGAP1* and *ZNF774 genes*, although it did not reach genome-wide significance.

Secondary analyses revealed five TWAS-associated genes, some of which have already been linked to severe COVID-19. In a comprehensive multitissue gene expression profiling study^32^, decreased expression of *CAMP* and *S100A8*/*S100A9* genes in patients with severe COVID-19 was observed, while another study detected the upregulation of *SCL25A37* among patients with severe COVID-19^33^. *SMARCC1* is a subunit of the SWI/SNF chromatin remodeling complex that has been identified as proviral for SARS-CoV-2 and other coronavirus strains through a genome-wide screen^34^. This complex is crucial for *ACE2* expression and viral entry into the cell^35^. However, it should be noted that using eQTL mostly from European populations such as those in GTEx could result in reduced power to detect associations.

To explore the genetic architecture of the trait among admixed AMR populations, we performed two cross-ancestry meta-analyses including the SCOURGE Latin American cohort GWAS findings. We found that the two novel risk variants were not associated with COVID-19 hospitalization outside the population-specific meta-analysis, highlighting the importance of complementing trans-ancestry meta-analyses with group-specific analyses. Notably, this analysis did not replicate the association at the *DSTYK* locus, which was associated with severe COVID-19 in Brazilian individuals with higher European admixture^36^. This lack of replication aligns with the initial hypothesis of that study suggesting that the risk haplotype was derived from European populations, as we reduced the weight of this ancestral contribution in our study by excluding those individuals.

Moreover, these cross-ancestry meta-analyses pointed to three loci that were not genome-wide significant in the HGIv7 ALL meta-analysis: a novel locus at *CREBBP* and two loci at *ZBTB7A* and *CASC20* that were reported in another meta-analysis. *CREBBP* and *ZBTB7A* achieved a stronger significance when considering only the EUR, AFR, and admixed AMR GIA groups. According to a recent study, elevated levels of the *ZBTB7A* gene promote a quasihomeostatic state between coronaviruses and host cells, preventing cell death by regulating oxidative stress pathways^37^. This gene is involved in several signaling pathways, such as B and T-cell differentiation^38^. On a separate note, *CREBBP* encodes the CREB binding protein (CBP), which is involved in transcriptional activation and is known to positively regulate the type I interferon response through virus-induced phosphorylation of IRF-3^39^. In addition, the CREBP/CBP interaction has been implicated in SARS-CoV-2 infection^40^ via the cAMP/PKA pathway. In fact, cells with suppressed *CREBBP* gene expression exhibit reduced replication of the so-called Delta and Omicron SARS-CoV-2 variants^40^.

We developed a cross-population PGS model, which effectively stratified individuals based on their genetic risk and demonstrated consistency with the clinical severity classification of the patients. Only a few polygenic scores were derived from COVID-19 GWAS data. Horowitz et al. (2022)^41^ developed a score using 6 and 12 associated variants (PGS ID: PGP000302) and reported an associated OR (top 10% vs rest) of 1.38 for risk of hospitalization in European populations, whereas the OR for Latin-American populations was 1.56. Since their sample size and the number of variants included in the PGS were lower, direct comparisons are not straightforward. Nevertheless, our analysis provides the first results for a PRS applied to a relatively large AMR cohort, being of value for future analyses regarding PRS transferability.

This study is subject to limitations, mostly concerning sample recruitment and composition. The SCOURGE Latino-American sample size is small, and the GWAS is likely underpowered. Another limitation is the difference in case □ control recruitment across sampling regions that, yet controlled for, may reduce the ability to observe significant associations driven by different compositions of the populations. In this sense, the identified risk loci might not replicate in a cohort lacking any of the parental population sources from the three-way admixture. Likewise, we could not explicitly control for socioenvironmental factors that could have affected COVID-19 spread and hospitalization rates, although genetic principal components are known to capture nongenetic factors. Finally, we must acknowledge the lack of a replication cohort. We used all the available GWAS data for COVID-19 hospitalization in admixed AMR in this meta-analysis due to the low number of studies conducted. Therefore, we had no studies to replicate or validate the results. These concerns may be addressed in the future by including more AMR GWAS in the meta-analysis, both by involving diverse populations in study designs and by supporting research from countries in Latin America.

This study provides novel insights into the genetic basis of COVID-19 severity, emphasizing the importance of considering host genetic factors by using non-European populations, especially of admixed sources. Such complementary efforts can pin down new variants and increase our knowledge on the host genetic factors of severe COVID-19.

## Materials and methods

### GWAS in Latin Americans from SCOURGE

#### The SCOURGE Latin American cohort

A total of 3,729 COVID-19-positive cases were recruited across five countries from Latin America (Mexico, Brazil, Colombia, Paraguay, and Ecuador) by 13 participating centers (supplementary Table 1) from March 2020 to July 2021. In addition, we included 1,082 COVID-19-positive individuals recruited between March and December 2020 in Spain who either had evidence of origin from a Latin American country or showed inferred genetic admixture between AMR, EUR, and AFR (with < 0.05% SAS/EAS). These individuals were excluded from a previous SCOURGE study that focused on participants with European genetic ancestries^16^. We used hospitalization as a proxy for disease severity and defined COVID-19-positive patients who underwent hospitalization as a consequence of the infection as cases and those who did not need hospitalization due to COVID-19 as controls.

Samples and data were collected with informed consent after the approval of the Ethics and Scientific Committees from the participating centers and by the Galician Ethics Committee Ref 2020/197. Recruitment of patients from IMSS (in Mexico, City) was approved by the National Committee of Clinical Research from Instituto Mexicano del Seguro Social, Mexico (protocol R-2020-785-082).

Samples and data were processed following normalized procedures. The REDCap electronic data capture tool^42,43^, hosted at Centro de Investigación Biomédica en Red (CIBER) from the Instituto de Salud Carlos III (ISCIII), was used to collect and manage demographic, epidemiological, and clinical variables. Subjects were diagnosed with COVID-19 based on quantitative PCR tests (79.3%) or according to clinical (2.2%) or laboratory procedures (antibody tests: 16.3%; other microbiological tests: 2.2%).

#### SNP array genotyping

Genomic DNA was obtained from peripheral blood and isolated using the Chemagic DNA Blood 100 kit (PerkinElmer Chemagen Technologies GmbH), following the manufacturer’s recommendations.

Samples were genotyped with the Axiom Spain Biobank Array (Thermo Fisher Scientific) following the manufacturer’s instructions in the Santiago de Compostela Node of the National Genotyping Center (CeGen-ISCIII; http://www.usc.es/cegen). This array contains probes for genotyping a total of 757,836 SNPs. Clustering and genotype calling were performed using Axiom Analysis Suite v4.0.3.3 software.

#### Quality control steps and variant imputation

A quality control (QC) procedure using PLINK 1.9^44^ was applied to both samples and the genotyped SNPs. We excluded variants with a minor allele frequency (MAF) <1%, a call rate <98%, and markers strongly deviating from Hardy-Weinberg equilibrium expectations (*p*<1×10^−6^) with mid-p adjustment. We also explored the excess of heterozygosity to discard potential cross-sample contamination. Samples missing >2% of the variants were filtered out. Subsequently, we kept the autosomal SNPs, removed high-LD regions and conducted LD pruning (windows of 1,000 SNPs, with a step size of 80 and a r^2^ threshold of 0.1) to assess kinship and estimate the global ancestral proportions. Kinship was evaluated based on IBD values, removing one individual from each pair with PI_HAT>0.25 that showed a Z0, Z1, and Z2 coherent pattern (according to the theoretical expected values for each relatedness level). Genetic principal components (PCs) were calculated with PLINK with the subset of LD pruned variants.

Genotypes were imputed with the TOPMed version r2 reference panel (GRCh38) using the TOPMed Imputation Server, and variants with Rsq<0.3 or with MAF<1% were filtered out. A total of 4,348 individuals and 10,671,028 genetic variants were included in the analyses.

#### Genetic admixture estimation

Global genetic inferred ancestry (GIA), referred to the genetic similarity to the used reference individuals, was estimated with ADMIXTURE^45^ v1.3 software following a two-step procedure. First, we randomly sampled 79 European (EUR) and 79 African (AFR) samples from The 1000 Genomes Project (1KGP)^46^ and merged them with the 79 Native American (AMR) samples from Mao *et al*.^47^ keeping the biallelic SNPs. LD-pruned variants were selected from this merge using the same parameters as in the QC. We then run an unsupervised analysis with K=3 to redefine and homogenize the clusters and to compose a refined reference for the analyses by applying a threshold of ≥95% of belonging to a particular cluster. As a result, 20 AFR, 18 EUR, and 38 AMR individuals were removed. The same LD-pruned variants data from the remaining individuals were merged with the SCOURGE Latin American cohort to perform supervised clustering and estimate admixture proportions. A total of 471 samples from the SCOURGE cohort with >80% estimated European GIA were removed to reduce the weight of the European ancestral component, leaving a total of 3,512 admixed Latin American (AMR) subjects for downstream analyses.

#### Association analysis

The results for the SCOURGE Latin American GWAS were obtained by testing for COVID-19 hospitalization as a surrogate of severity. To accommodate the continuum of GIA in the cohort, we opted for a joint testing of all the individuals as a single study using a mixed regression model, as this approach has demonstrated a greater power and sufficient control of population structure^48^. The SCOURGE cohort consisted of 3,512 COVID-19-positive patients: cases (n=1,625) were defined as hospitalized COVID-19 patients, and controls (n=1,887) were defined as nonhospitalized COVID-19-positive patients.

Logistic mixed regression models were fitted using the SAIGEgds^49^ package in R, which implements the two-step mixed SAIGE^50^ model methodology and the SPA test. Baseline covariables included sex, age (continuous), and the first 10 PCs. To account for potential heterogeneity in the recruitment and hospitalization criteria across the participating countries, we adjusted the models by groups of the recruitment areas classified into six categories: Brazil, Colombia, Ecuador, Mexico, Paraguay, and Spain. This dataset has not been used in any previously published GWAS of COVID-19.

### Meta-analysis of Latin American populations

The results of the SCOURGE Latin American cohort were meta-analyzed with the AMR HGI-B2 data, conforming our primary analysis. Summary results from the HGI freeze 7 B2 analysis corresponding to the admixed AMR population were obtained from the public repository (April 8, 2022: https://www.covid19hg.org/results/r7/), summing up 3,077 cases and 66,686 controls from seven contributing studies. We selected the B2 phenotype definition because it offered more power, and the presence of population controls not ascertained for COVID-19 does not have a drastic impact on the association results.

The meta-analysis was performed using an inverse-variance weighting method in METAL^51^. The average allele frequency was calculated, and variants with low imputation quality (Rsq<0.3) were filtered out, leaving 10,121,172 variants for meta-analysis.

Heterogeneity between studies was evaluated with Cochran’s Q test. The inflation of results was assessed based on a genomic control (lambda).

#### Replicability of associations

The model-based method MAMBA^52^ was used to calculate the posterior probabilities of replication for each of the lead variants. Variants with p<1×10^−05^ were clumped and combined with random pruned variants from the 1KGP AMR reference panel. Then, MAMBA was applied to the set of significant and non-significant variants.

Each of the lead variants was also tested for association with the main comorbidities in the SCOURGE cohort with logistic regression models (adjusted by the same base covariables as the GWAS).

### Definition of the genetic risk loci and putative functional impact

#### Definition of lead variant and novel loci

To define the lead variants in the loci that were genome-wide significant, LD-clumping was performed on the meta-analysis data using a threshold p-value<5×10^−8^, clump distance=1500 kb, independence set at a threshold r^2^=0.1 and the SCOURGE cohort genotype data as the LD reference panel. Independent loci were deemed as a novel finding if they met the following criteria: 1) p-value<5×10^−8^ in the meta-analysis and p-value>5×10^−8^ in the HGI B2 ALL meta-analysis or in the HGI B2 AMR and AFR and EUR analyses when considered separately; 2) Cochran’s Q-test for heterogeneity of effects is <0.05/N_loci_, where N_loci_ is the number of independent variants with p<5×10^−8^; and 3) the nearest gene has not been previously described in the latest HGIv7 update.

#### Annotation and initial mapping

Functional annotation was performed with FUMA^53^ for those variants with a p-value<5×10^−8^ or in moderate-to-strong LD (r^2^>0.6) with the lead variants, where the LD was calculated from the 1KGP AMR panel. Genetic risk loci were defined by collapsing LD blocks within 250 kb. Then, genes, scaled CADD v1.4 scores, and RegulomeDB v1.1 scores were annotated for the resulting variants with ANNOVAR in FUMA^53^. Gene-based analysis was also performed using MAGMA^54^ as implemented in FUMA under the SNP-wide mean model using the 1KGP AMR reference panel. Significance was set at a threshold p<2.66×10^−6^ (which assumes that variants can be mapped to a total of 18,817 genes).

FUMA allowed us to perform initial gene mapping by two approaches: (1) positional mapping, which assigns variants to genes by physical distance using 10-kb windows; and (2) eQTL mapping based on GTEx v.8 data from whole blood, lungs, lymphocytes, and esophageal mucosa tissues, establishing a false discovery rate (FDR) of 0.05 to declare significance for variant-gene pairs.

Subsequently, to assign the variants to the most likely gene driving the association, we refined the candidate genes by fine mapping the discovered regions

#### Bayesian fine-mapping

To conduct a Bayesian fine mapping, credible sets for the genetic loci considered novel findings were calculated on the results from each of the three meta-analyses to identify a subset of variants most likely containing the causal variant at the 95% confidence level, assuming that there is a single causal variant and that it has been tested. We used *corrcoverage* (https://cran.rstudio.com/web/packages/corrcoverage/index.html) for R to calculate the posterior probabilities of the variant being causal for all variants with a r^2^>0.1 with the leading SNP and within 1 Mb except for the novel variant in chromosome 19, for which we used a window of 0.5 Mb. Variants were added to the credible set until the sum of the posterior probabilities was ≥0.95.

#### VEP and V2G annotation

We used the Variant-to-Gene (V2G) score to prioritize the genes that were most likely affected by the functional evidence based on expression quantitative trait loci (eQTL), chromatin interactions, in silico functional predictions, and distance between the prioritized variants and transcription start site (TSS), based on data from the Open Targets Genetics portal^55^. Details of the data integration and the weighting of each of the datasets are described with detail here: https://genetics-docs.opentargets.org/our-approach/data-pipeline. V2G is a score for ranking the functional genomics evidence that supports the connection between variants and genes (the higher the score the more likely the variant to be functionally implicated on the assigned gene). We used VEP release 111 (URL: https://www.ensembl.org/info/docs/tools/vep/index.html; accessed April 10, 2024) ^56^ to annotate the following: gene symbol, function (exonic, intronic, intergenic, non-coding RNA, etc.), impact, feature type, feature, and biotype.

We queried the GWAS Catalog (date of accession: 01/07/2024) for evidence of association of each of the prioritized genes with traits related to lung diseases or phenotypes. Lastly, those which were linked to COVID-19, infection, or lung diseases in the revised literature were classified as “literature evidence”.

### Transcription-wide association studies

Transcriptome-wide association studies (TWAS) were conducted using the pretrained prediction models with MASHR-computed effect sizes on GTEx v8 datasets^57,58^. The results from the Latin American meta-analysis were harmonized and integrated with the prediction models through S-PrediXcan^59^ for lung, whole blood, lymphocyte and esophageal mucosal tissues. Statistical significance was set at p-value<0.05 divided by the number of genes that were tested for each tissue. Subsequently, we leveraged results for all 49 tissues and ran a multitissue TWAS to improve the power for association, as demonstrated recently^60^. TWAS was also performed using recently published gene expression datasets derived from a cohort of African Americans, Puerto Ricans, and Mexican Americans (GALA II-SAGE)^61^.

### Cross-population meta-analyses

We conducted two additional meta-analyses to investigate the ability of combining populations to replicate our discovered risk loci. This methodology enabled the comparison of effects and the significance of associations in the novel risk loci between the results from analyses that included or excluded other population groups.

The first meta-analysis comprised the five populations analyzed within HGI (B2-ALL). Additionally, to evaluate the three GIA components within the SCOURGE Latin American cohort^62^, we conducted a meta-analysis of the admixed AMR, EUR, and AFR cohorts (B2). All summary statistics were retrieved from the HGI repository. We applied the same meta-analysis methodology and filters as in the admixed AMR meta-analysis.

### Cross-population Polygenic Risk Score

A polygenic risk score (PGS) for critical COVID-19 was derived by combining the variants associated with hospitalization or disease severity that have been discovered to date. We curated a list of lead variants that were 1) associated with either severe disease or hospitalization in the latest HGIv7 release^1^ (using the hospitalization weights) or 2) associated with severe disease in the latest GenOMICC meta-analysis^2^ that were not reported in the latest HGI release. A total of 48 markers were used in the PGS model (see supplementary Table 13) since two variants were absent from our study.

Scores were calculated and normalized for the SCOURGE Latin American cohort with PLINK 1.9. This cross-ancestry PGS was used as a predictor for hospitalization (COVID-19-positive patients who were hospitalized *vs*. COVID-19-positive patients who did not necessitate hospital admission) by fitting a logistic regression model. Prediction accuracy for the PGS was assessed by performing 500 bootstrap resamples of the increase in the pseudo-R-squared. We also divided the sample into deciles and percentiles to assess risk stratification. The models were fit for the dependent variable adjusting for sex, age, the first 10 PCs, and the sampling region (in the Admixed AMR cohort) with and without the PGS, and the partial pseudo-R2 was computed and averaged among the resamples.

A clinical severity scale was used in a multinomial regression model to further evaluate the power of this cross-ancestry PGS for risk stratification. These severity strata were defined as follows: 0) asymptomatic; 1) mild, that is, with symptoms, but without pulmonary infiltrates or need of oxygen therapy; 2) moderate, that is, with pulmonary infiltrates affecting <50% of the lungs or need of supplemental oxygen therapy; 3) severe disease, that is, with hospital admission and PaO_2_<65 mmHg or SaO_2_<90%, PaO_2_/FiO_2_<300, SaO_2_/FiO_2_<440, dyspnea, respiratory frequency≥22 bpm, and infiltrates affecting >50% of the lungs; and 4) critical disease, that is, with an admission to the ICU or need of mechanical ventilation (invasive or noninvasive).

## Data Availability

Summary statistics from the SCOURGE Latin-American GWAS will be available at https://github.com/CIBERER/Scourge-COVID19

## Data availability

Summary statistics from the SCOURGE Latin American GWAS and the analysis scripts are available from the public repository https://github.com/CIBERER/Scourge-COVID19.

## Author contributions

Study design: RC, AC, CF. Data collection: SCOURGE cohort group. Data analysis: SD-DA, RC, ADL, CF, JML-S. Interpretation: SD-DA, RC, ADL. Drafting of the manuscript: SD-DA, RC, ADL, CF, AR-M, AC. Critical revision of the manuscript: SD-DA, RC, ADL, AC, CF, JAR, AR-M, and PL. Approval of the final version of the publication: all coauthors.

## Acknowledgments

The contribution of the Centro National de Genotipado (CEGEN) and Centro de Supercomputación de Galicia (CESGA) for funding this project by providing supercomputing infrastructures is also acknowledged. The authors are also particularly grateful for the supply of material and the collaboration of patients, health professionals from participating centers and biobanks. Namely, Biobanc-Mur, and biobancs of the Complexo Hospitalario Universitario de A Coruña, Complexo Hospitalario Universitario de Santiago, Hospital Clínico San Carlos, Hospital La Fe, Hospital Universitario Puerta de Hierro Majadahonda—Instituto de Investigación Sanitaria Puerta de Hierro—Segovia de Arana, Hospital Ramón y Cajal, IDIBGI, IdISBa, IIS Biocruces Bizkaia, IIS Galicia Sur. Also biobanks of the Sistema de Salud de Aragón, Sistema Sanitario Público de Andalucía, and Banco Nacional de ADN.

