## Supplementary Material for "Novel risk loci for COVID-19 hospitalization among admixed American populations"

**Supplemental figures**

**Supplemental Figure 1. Global Genetic Inferred Ancestry (GIA) composition in the SCOURGE Latin-American cohort.** European (EUR), African (AFR) and Native American (AMR) GIA was derived with ADMIXTURE from a reference panel composed of Aymaran, Mayan, Nahuan, and Quechuan individuals of Native-American genetic ancestry and randomly selected samples from the EUR and AFR 1KGP populations. The colours represent the different geographical sampling regions from which the admixed American individuals from SCOURGE were recruited.

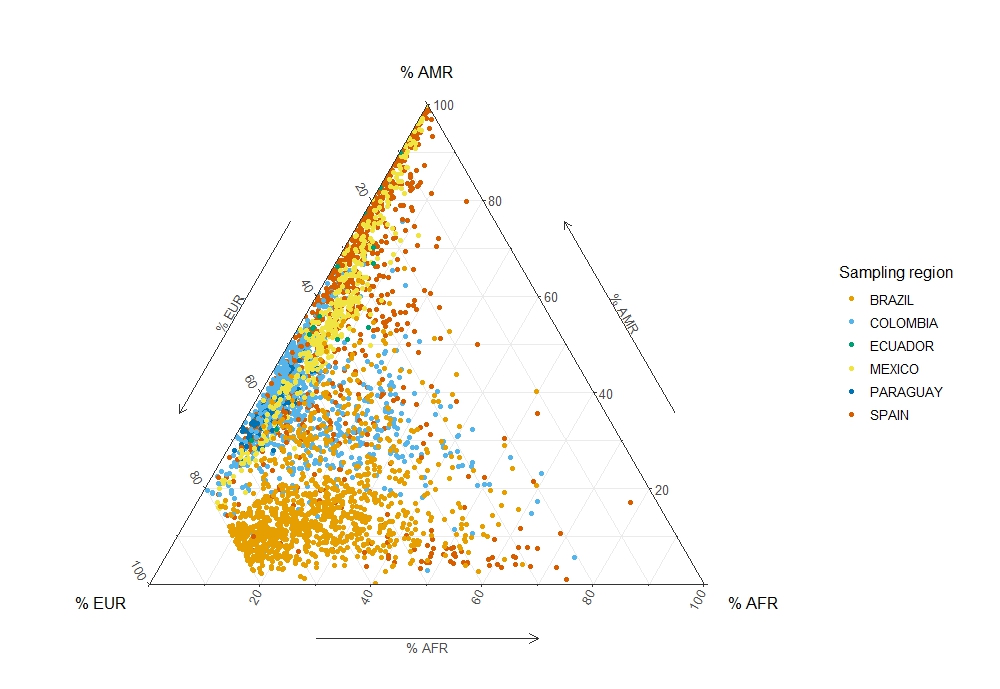

**Supplemental Figure 2. Quantile-Quantile plot for the AMR GWAS meta-analysis.** A lambda inflation factor of 1.015 was obtained.

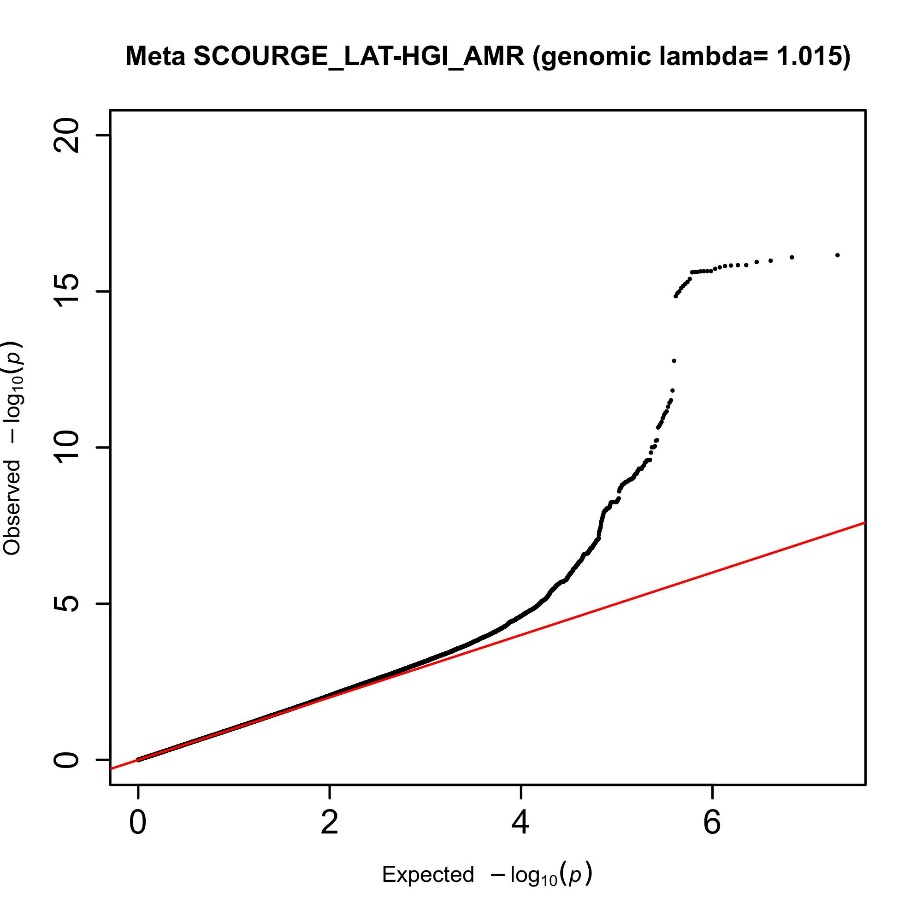

**Supplemental Figure 3. Regional association plots for the fine mapped loci in chromosomes 2 (upper panel) and 16 (lower panel).** Coloured in red, the variants allocated to the credible set at the 95% confidence according to the Bayesian fine mapping. In blue, the sentinel variant.

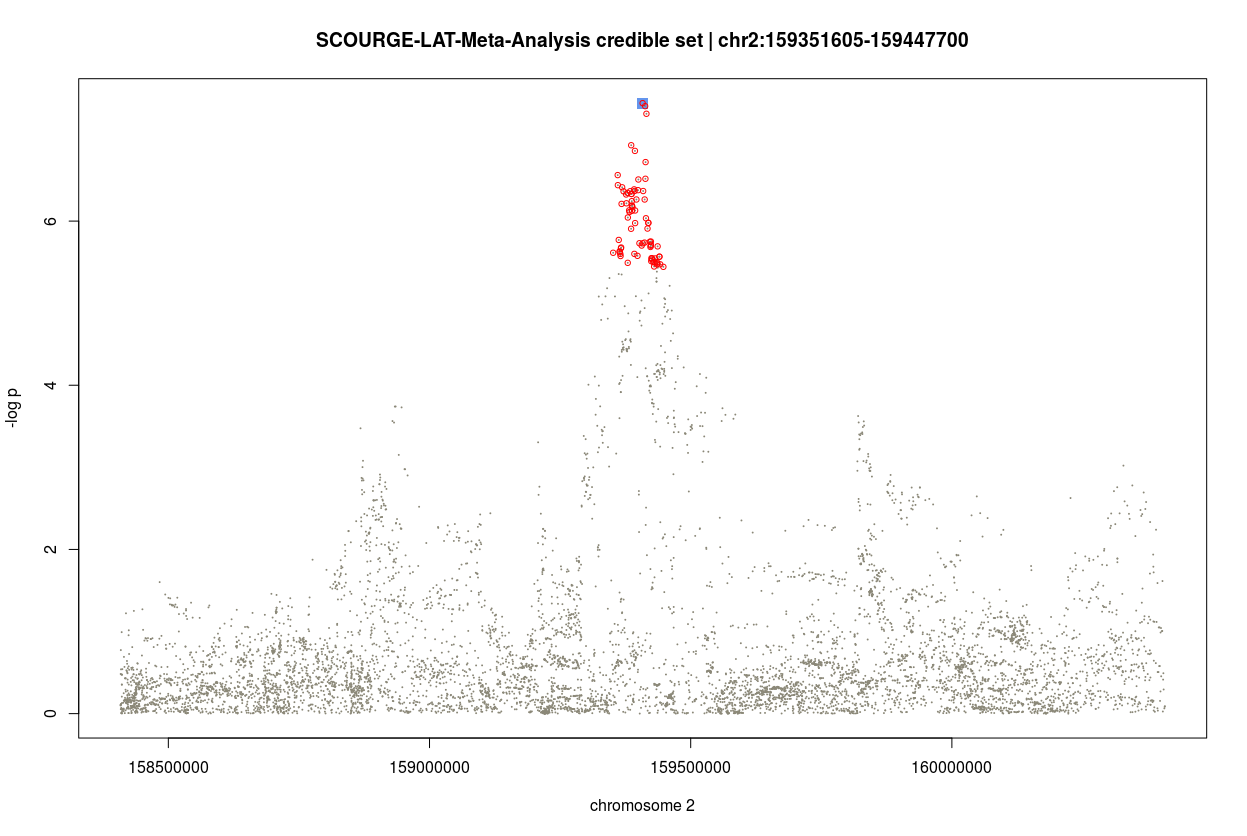

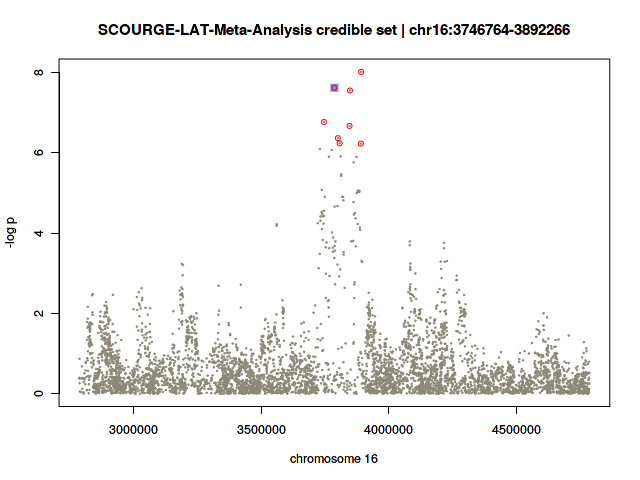

**Supplemental Figure 4. Gene-tissue pairs for which either rs1003835 or rs60606421 are significant eQTLs at FDR<0.05 (data retrieved from https://gtexportal.org/home/snp/).** rs1003835 (chromosome 2) maps to *BAZ2B*, *LY75*, and *PLA2R1* genes. As for the lead variant of chromosome 11, rs77599934, since it was not an eQTL, we used an LD proxy variant (rs60606421). *DDIAS* and *PRCP* genes map closely to this variant. NES and p-values correspond to the normalized effect size (and direction) of eQTL-gene associations and the p-value for the tissue, respectively.

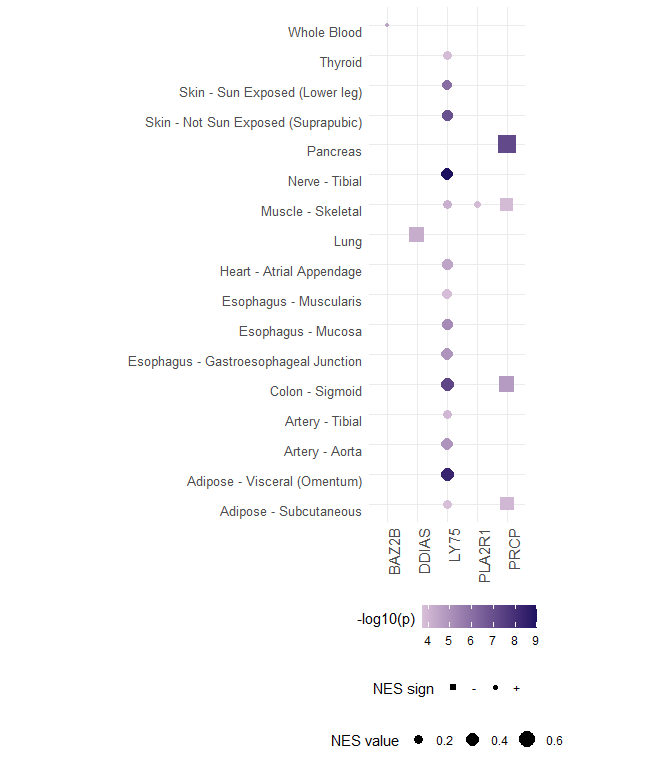

**Full list of cohort members and affiliations**

### Scourge Cohort Group

Javier Abellan^1,2^; René Acosta-Isaac^3^; Jose María Aguado^4,5,6,7^; Carlos Aguilar^8^; Sergio Aguilera-Albesa^9,10^; Abdolah Ahmadi Sabbagh^11^; Jorge Alba^12^; Sergiu Albu^13,14,15^; Karla A.M. Alcalá-Gallardo^16^; Julia Alcoba-Florez^17^; Sergio Alcolea Batres^18^; Holmes Rafael Algarin-Lara^19,20^; Virginia Almadana^21^; Julia Almeida^22,23,24,25^; Berta Almoguera^26,27^; María R. Alonso^28^; Nuria Alvarez^28^; Rodolfo Alvarez-Sala Walther^18^; Mónica T. Andrade ^29,30^; Álvaro Andreu-Bernabeu^31,6^; Maria Rosa Antonijoan^32^; Eunate Arana-Arri^33,34^; Carlos Aranda^35,36^; Celso Arango^31,37,6^; Carolina Araque^38,39^; Nathalia K. Araujo^40^; Izabel M.T. Araujo^41^; Ana C. Arcanjo^42,43,44^; Ana Arnaiz^45,46,47^; Francisco Arnalich Fernández^48^; María J. Arranz^49^; José Ramon Arribas Lopez^48^; Maria-Jesus Artiga^50^; Yubelly Avello-Malaver^51^; Carmen Ayuso^26,27^; Ana Margarita Baldión^51^; Belén Ballina Martín^11^; Raúl C. Baptista-Rosas^52,53,54^; Andrea Barranco-Díaz^20^; María Barreda- Sánchez^55,56^; Viviana Barrera-Penagos^51^; Moncef Belhassen-Garcia^57,58^; Enrique Bernal^55^; David Bernal-Bello^59^; Joao F. Bezerra^60^; Marcos A.C. Bezerra^61^; Natalia Blanca-López^62^; Rafael Blancas^63^; Lucía Boix-Palop^64^; Alberto Borobia^65^; Elsa Bravo^66^; María Brion^67,68^; Óscar Brochado-Kith^69,7^; Ramón Brugada^70,71,68,72^; Matilde Bustos^73^; Alfonso Cabello^74^; Juan J. Caceres-Agra^75^; Esther Calbo^76^; Enrique J. Calderón^77,78,79^; Shirley Camacho^80^; Cristina Carbonell^81,58^; Servando Cardona-Huerta^82^; Antonio Augusto F. Carioca^83^; Maria Sanchez Carpintero^35,36^; Carlos Carpio Segura^18^; Thássia M.T. Carratto^84^; José Antonio Carrillo-Avila^85^; Maria C.C. Carvalho^86^; Carlos Casasnovas^87,88,27^; Luis Castano^33,34,27,89,90^; Carlos F. Castaño^35,36^; Jose E. Castelao^91^; Aranzazu Castellano Candalija^92^; María A. Castillo^80^; Yolanda Cañadas^36^; Francisco C. Ceballos^27^; Jessica G. Chaux^39^; Walter G. Chaves- Santiago^93,39^; Sylena Chiquillo-Gómez^19,20^; Marco A. Cid-Lopez^16^; Oscar Cienfuegos-Jimenez^82^; Rosa Conde-Vicente^94^; M. Lourdes Cordero-Lorenzana^95^; Dolores Corella^96,97^; Almudena Corrales^98,99^; Jose L. Cortes-Sanchez^82,100^; Marta Corton^26,27^; Tatiana X. Costa^101^; Raquel Cruz^27,102^; Marina S. Cruz^40^; Luisa Cuesta^103^; Gabriela C.R. Cunha^104^; David Dalmau^105,76^; Raquel C.S. Dantas-Komatsu^40^; M. Teresa Darnaude^106^; Alba De Martino-Rodríguez^107,108^; Juan De la Cruz Troca^109,110,78^; Juan Delgado-Cuesta^111^; Aranzazu Diaz de Bustamante^106^; Covadonga M. Diaz-Caneja^31,37,6^; Beatriz Dietl^76^; Silvia Diz-de Almeida^27,102^; Elena Domínguez-Garrido^112^; Alice M. Duarte^41^; Anderson Díaz-Pérez^20^; Jose Echave-Sustaeta^113^; Rocío Eiros^114^; César O. Enciso-Olivera^38,39^; Gabriela Escudero^115^; Pedro Pablo España^116^; Gladys Mercedes Estigarribia Sanabria^117^; María Carmen Fariñas^45,46,47^; Marianne R. Fernandes^118,119^; Lidia Fernandez-Caballero^26,27^; María J. Fernandez-Nestosa^120^; Ramón Fernández^45,121^; Silvia Fernández Ferrero^11^; Yolanda Fernández Martínez^11^; Ana Fernández-Cruz^122^; Uxía Fernández-Robelo^123^; Amanda Fernández-Rodríguez^69,7^; Marta Fernández-Sampedro^45,47,46^; Ruth Fernández-Sánchez^26,27^; Tania Fernández-Villa^124,78^; Carmen Fernéndez Capitán^92^; Patricia Flores-Pérez^125^; Vicente Friaza^78,79^; Lácides Fuenmayor-Hernández^20^; Marta Fuertes Núñez^11^; Victoria Fumadó^126^; Ignacio Gadea^127^; Lidia Gagliardi^35,36^; Manuela Gago-Domínguez^128,129^; Natalia Gallego^130^; Cristina Galoppo^131^; Carlos Garcia-Cerrada^1,2,132^; Josefina Garcia-García^55^; Inés García^26,27^; Mercedes García^35,36^; Leticia García^35,36^; María Carmen García Torrejón^133,2^; Irene García-García^65^; Carmen García-Ibarbia^45,47,46^; Andrés C. García-Montero^134^; Ana García-Soidán^135^; Elisa García-Vázquez^55^; Aitor García-de-Vicuña^33,136^; Emiliano Garza-Frias^82^; Jesus Gaytán-Martínez^137^, Angela Gentile^131^; Belén Gil-Fournier^138^; Fernan Gonzalez Bernaldo de Quirós^139^; Manuel Gonzalez-Sagrado^94^; Hugo Gonzalo Benito^140^; Beatriz González Álvarez^107,108^; Anna González-Neira^28^; Javier González-Peñas^31,6,37^; Oscar Gorgojo-Galindo^141^; Florencia Guaragna^131^; Genilson P. Guegel^142^; Beatriz Guillen-Guio^98^; Encarna Guillen-Navarro^55,143,144,27^; Pablo Guisado-Vasco^113^; Luz D. Gutierrez-Castañeda^145,39^; Juan F. Gutiérrez-Bautista^146^; Luis Gómez Carrera^18^; María Gómez García^128^; Ángela Gómez Sacristán^147^; Javier Gómez-Arrue^107,108^; Mario Gómez-Duque^93,39^; Miguel Górgolas^74^; Sarah Heili-Frades^148^; Estefania Hernandez^149^; Luis D. Hernandez-Ortega^150,151^; Cristina Hernández Moro^11^; Guillermo Hernández-Pérez^81^; Rebeca Hernández-Vaquero^152^; Belen Herraez^28^; M. Teresa Herranz^55^; María Herrera^35,36^; María José Herrero^153,154^; Antonio Herrero-Gonzalez^155^; Juan P. Horcajada^156,157,14,158,7^; Natale Imaz-Ayo^33^; Maider Intxausti-Urrutibeaskoa^159^; Rafael H. Jacomo^160^; Rubén Jara^55^; Perez Maria Jazmin^131^; María A. Jimenez-Sousa^69,7^; Ángel Jiménez^35,36^; Pilar Jiménez^146^; Ignacio Jiménez-Alfaro^161^; Iolanda Jordan^162,163,78^; Rocío Laguna-Goya^164,165^; Daniel Laorden^18^; María Lasa-Lazaro^164,165^; María Claudia Lattig^80,166^; Ailen Lauriente^131^; Anabel Liger Borja^167^; Lucía Llanos^169^; Esther Lopez-Garcia^109,110,78,170^; Rosario Lopez-Rodriguez^26,27^; Leonardo Lorente^171^; José E. Lozano^172^; María Lozano-Espinosa^167^; Andre D. Luchessi^173^; Eduardo López Granados^174,175,27^; Amparo López-Bernús^81,58^; Miguel A. López-Ruz^176,177,178^; Aluísio X. Magalhães-Brasil ^179^;Ignacio Mahillo^180,181,99^; Esther Mancebo^164,165^; Carmen Mar^116^; Cristina Marcelo Calvo^92^; Miguel Marcos^81,58^; Alba Marcos-Delgado^124^; Pablo Mariscal Aguilar^18^; Marta Martin-Fernandez^182^; Laura Martin-Pedraza^62^; Amalia Martinez^183^; Iciar Martinez-Lopez^184,185^; Oscar Martinez-Nieto^51,166^; Pedro Martinez-Paz^140^; Angel Martinez-Perez^186^; Michel F. Martinez-Resendez^82^; María M. Martín^187^; María Dolores Martín^188^; Vicente Martín^124,78^; Caridad Martín-López^167^; José-Ángel Martín-Oterino^81,58^; María Martín-Vicente^69^; Ricardo Martínez^149^; Juan José Martínez^88,27^; Silvia Martínez^45,47^; Violeta Martínez Robles^11^; Eleno Martínez-Aquino^189^; Óscar Martínez-González^190^; Andrea Martínez-Ramas^26,27^; Laura Marzal^26,27^; Alicia Marín Candon^65^; Jose Antonio Mata-Marin,^137^ Juliana F. Mazzeu^179,191,192^; Jeane F.P. Medeiros^40^; Francisco J. Medrano^77,78,79^; Xose M. Meijome^193,194^; Natalia Mejuto-Montero^195^; Celso T. Mendes-Junior ^84,196,197^; Humberto Mendoza Charris^66,20^; Eleuterio Merayo Macías^198^; Fátima Mercadillo^199^; Arieh R. Mercado-Sesma^150,151^; Pablo Minguez^26,27^; Antonio J J. Molina^124,78^; Elena Molina-Roldán^200^; Juan José Montoya^149^; Patricia Moreira-Escriche^201^; Xenia Morelos-Arnedo^66,20^; Victor Moreno Cuerda^1,2^; Alberto Moreno Fernández^92^; Antonio Moreno-Docón^55^; Junior Moreno-Escalante^20^; Rubén Morilla^79,202^; Patricia Muñoz García^203,99,6^; Ana Méndez-Echevarria^204^; Pablo Neira^131^; Julian Nevado^27,131,205^; Israel Nieto-Gañán^135^; Joana F.R. Nunes^42^; Rocio Nuñez- Torres^28^; Antònia Obrador-Hevia^206,207^; J. Gonzalo Ocejo-Vinyals^45,47^; Virginia Olivar^131^; Silviene F. Oliveira^179,208,208,210,211^; Lorena Ondo^26,27^; Alberto Orfao^22,23,24,25^; Luis Ortega^212^; Eva Ortega-Paino^50^; Fernando Ortiz-Flores^45,47^; Rocio Ortiz-Lopez^213,82^; José A. Oteo^12,214^; Harry Pachajoa^215,216^; Manuel Pacheco^149^; Fredy Javier Pacheco-Miranda^20^; Irene Padilla Conejo^11^; Sonia Panadero-Fajardo^85^; Mara Parellada^31,37,6^; Roberto Pariente-Rodríguez^131^; Estela Paz-Artal^164,165,217^; Germán Peces-Barba^218,99^; Miguel S. Pedromingo Kus^219^; Celia Perales^127^; Patricia Perez^220^; Gustavo Perez-de-Nanclares^33,221^; Teresa Perucho^222^; Aline Pic-Taylor ^42,208,209,211^, Lisbeth A. Pichardo^11^; Mel·lina Pinsach-Abuin^70,68^; Luz Adriana Pinzón^93,39^; Guillermo Pita^30^; Francesc Pla-Junca^223,27^; Laura Planas-Serra^88,27^; Ericka N. Pompa-Mera^224,137^ ; Gloria L. Porras-Hurtado^149^; Aurora Pujol^88,27,225^; César Pérez^226^; Felipe Pérez-García^227,228^; Patricia Pérez-Matute^214^; Alexandra Pérez-Serra^70,68^; M. Elena Pérez-Tomás^55^; María Eugenia Quevedo Chávez^19,20^; Maria Angeles Quijada^30,229^; Inés Quintela^128^; Diana Ramirez-Montaño^230^; Soraya Ramiro León^131^; Pedro Rascado Sedes^231^; Delia Recalde^107,108^; Emma Recio-Fernández^214^; Salvador Resino^69,7^; Adriana P. Ribeiro^29,30,232^; Carlos S. Rivadeneira-Chamorro^39^; Diana Roa-Agudelo^51^; Montserrat Robelo Pardo^231^; Marilyn Johanna Rodriguez^39^; German Ezequiel Rodriguez Novoa^131^; Fernando Rodriguez-Artalejo^109,110,78,170^; Carlos Rodriguez-Gallego^233,234^; José A. Rodriguez-Garcia^11^; María A. Rodriguez-Hernandez^73^; Antonio Rodriguez-Nicolas^146^; Agustí Rodriguez-Palmero^235,88^; Paula A. Rodriguez-Urrego^51^; Belén Rodríguez Maya^1^; Marena Rodríguez-Ferrer^20^; Emilio Rodríguez-Ruiz^231,129^; Federico Rojo^236,25^; Andrea Romero-Coronado^20^; Filomeno Rondón García^11^; ~~Lidia S. Rosa~~^~~236~~^; Antonio Rosales-Castillo^237^; Cladelis Rubio^238,239^; María Rubio Olivera^35,36^; Montserrat Ruiz^88,27^; Francisco Ruiz-Cabello^146,177,240^; Eva Ruiz-Casares^222^; Juan J. Ruiz-Cubillan^45,47^; Javier Ruiz-Hornillos^241,36,242^; Pablo Ryan^243,244,245,7^; Hector D. Salamanca^38,39^; Lorena Salazar-García^80^; Giorgina Gabriela Salgueiro Origlia ^92^; Cristina Sancho-Sainz^159^; Jorge Luis Sandoval-Ramírez,^137^ Anna Sangil^64^; Arnoldo Santos^226^; Ney P.C. Santos^118^; Amanda C.M. Saúde ^30,246^ Agatha Schlüter^88,27^; Sonia Segovia^223,247,248^; Alex Serra-Llovich^249^; Fernando Sevil Puras^8^; Marta Sevilla Porras^27,130^; Miguel A. Sicolo^250,251^; Vivian N. Silbiger^173^; Nayara S. Silva^252^; ~~Fabiola T.C. Silva~~^~~40~~^~~;~~ Cristina Silván Fuentes^27^; Jordi Solé-Violán^253,99,254^; José Manuel Soria^186^; Jose V. Sorlí^96,97^; Renata R. Sousa^179^; Juan Carlos Souto^3^; Karla S.C. Souza^86^; Vanessa S. Souza^104^; John J. Sprockel^93,39^; David A. Suarez-Zamora^51^; José Javier Suárez-Rama^128^; Pedro-Luis Sánchez^114,58^; Antonio J. Sánchez López^255^; María Concepción Sánchez Prados^18^; Javier Sánchez Real^11^; Jorge Sánchez Redondo^1,256^; Clara Sánchez-Pablo^114^; Olga Sánchez-Pernaute^257^; Xiana Taboada-Fraga^195^; Eduardo Tamayo^258,140,7^; Alvaro Tamayo-Velasco^259^; Juan Carlos Taracido-Fernandez^155^; Nathali A.C. Tavares^260^; Carlos Tellería^107,108^; Jair Antonio Tenorio Castaño^27,130,205^; Alejandro Teper^131^; Ronald P. Torres Gutiérrez^221^; Juan Torres-Macho^261^; Lilian Torres-Tobar^39^; Jesús Troya^243^; Miguel Urioste^199^; Juan Valencia-Ramos^262^; Agustín Valido^21,263^; Juan Pablo Vargas Gallo^264,265^; Belén Varón^266^; Romero H.T. Vasconcelos^260^; Tomas Vega^267^; Santiago Velasco-Quirce^268^; Julia Vidán Estévez^11^; Miriam Vieitez-Santiago^45,47^; Carlos Vilches^269^; Lavinia Villalobos^11^; Felipe Villar^218^; Judit Villar-Garcia^270,271,272^; Cristina Villaverde^26,27^; Pablo Villoslada-Blanco^214^; Ana Virseda-Berdices^69^; Valentina Vélez-Santamaría^87,88^; Virginia Víctor^35,36^; Zuleima Yáñez^20^; Antonio Zapatero-Gaviria^273^; Ruth Zarate^274^; Sandra Zazo^236^; Gabriela V. da Silva^41^; Raimundo de Andrés^275^; Jéssica N.G. de Araújo^252^; Carmen de Juan^201^; Julianna Lys de Sousa Alves Neri^276^; Carmen de la Horra^79^; Ana B. de la Hoz^33^; Victor del Campo-Pérez^277^; Manoella do Monte Alves^278,279^; Katiusse A. dos Santos^86^; Yady Álvarez-Benítez^19,20^; Felipe Álvarez-Navia^81,58^; María Íñiguez^214^; Miguel López de Heredia^27^; Ingrid Mendes^27^; Rocío Moreno^27^; Esther Sande^27,129,102^; Carlos Flores^280,98,99,234^; José A. Riancho^45,46,47,27^; Augusto Rojas-Martinez^82^; Pablo Lapunzina^27,130,205^; Angel Carracedo^27,129,102,128^

#### Scourge Cohort Group’s filiations

^1^, Hospital Universitario Mostoles, Medicina Interna, Madrid, Spain

^2^, Universidad Francisco de Vitoria, Madrid, Spain

^3^, Haemostasis and Thrombosis Unit, Hospital de la Santa Creu i Sant Pau, IIB Sant Pau, Barcelona, Spain

^4^, Unit of Infectious Diseases, Hospital Universitario 12 de Octubre, Instituto de Investigación Sanitaria Hospital 12 de Octubre (imas12), Madrid, Spain

^5^, Spanish Network for Research in Infectious Diseases (REIPI RD16/0016/0002), Instituto de Salud Carlos III, Madrid, Spain

^6^, School of Medicine, Universidad Complutense, Madrid, Spain

^7^, Centro de Investigación Biomédica en Red de Enfermedades Infecciosas (CIBERINFEC), Instituto de Salud Carlos III, Madrid, Spain

^8^, Hospital General Santa Bárbara de Soria, Soria, Spain

^9^, Pediatric Neurology Unit, Department of Pediatrics, Navarra Health Service Hospital, Pamplona, Spain

^10^, Navarra Health Service, NavarraBioMed Research Group, Pamplona, Spain

^11^, Complejo Asistencial Universitario de León, León, Spain

^12^, Hospital Universitario San Pedro, Infectious Diseases Department, Logroño, Spain

^13^, Fundación Institut Guttmann, Institut Universitari de Neurorehabilitació adscrit a la UAB, Hospital de Neurorehabilitació, Barcelona, Spain

^14^, Universitat Autònoma de Barcelona (UAB), Barcelona, Spain

^15^, Fundació Institut d’Investigació en Ciències de la Salut Germans Trias i Pujol, Barcelona, Spain

^16^, Hospital General de Occidente, Guadalajara, Mexico

^17^, Microbiology Unit, Hospital Universitario Ntra. Sra. de Candelaria, Santa Cruz de Tenerife, Spain

^18^, Hospital Universitario La Paz-IDIPAZ, Servicio de Neumología, Madrid, Spain

^19^, Camino Universitario Adelita de Char, Mired IPS, Barranquilla, Colombia

^20^, Universidad Simón Bolívar, Facultad de Ciencias de la Salud, Barranquilla, Colombia

^21^, Hospital Universitario Virgen Macarena, Neumología, Seville, Spain

^22^, Departamento de Medicina, Universidad de Salamanca, Salamanca, Spain

^23^, Centro de Investigación del Cáncer (IBMCC) Universidad de Salamanca - CSIC, Salamanca, Spain

^24^, Biomedical Research Institute of Salamanca (IBSAL) Salamanca, Spain

^25^, Centre for Biomedical Network Research on Cancer (CIBERONC), Instituto de Salud Carlos III, Madrid, Spain

^26^, Department of Genetics & Genomics, Instituto de Investigación Sanitaria-Fundación Jiménez Díaz University Hospital - Universidad Autónoma de Madrid (IIS-FJD, UAM), Madrid, Spain

^27^, Centre for Biomedical Network Research on Rare Diseases (CIBERER), Instituto de Salud Carlos III, Madrid, Spain

^28^, Spanish National Cancer Research Centre, Human Genotyping-CEGEN Unit, Madrid, Spain

^29^, Hospital das Forças Armadas, Brazil

^30^, Exército Brasileiro, Brazil

^31^, Department of Child and Adolescent Psychiatry, Institute of Psychiatry and Mental Health, Hospital General Universitario Gregorio Marañón (IiSGM), Madrid, Spain

^32^, Clinical Pharmacology Service, Hospital de la Santa Creu i Sant Pau, IIB Sant Pau, Barcelona, Spain

^33^, Biocruces Bizkai HRI, Barakaldo, Bizkaia, Spain

^34^, Cruces University Hospital, Osakidetza, Barakaldo, Bizkaia, Spain

^35^, Hospital Infanta Elena, Valdemoro, Madrid, Spain

^36^, Instituto de Investigación Sanitaria-Fundación Jiménez Díaz University Hospital - Universidad Autónoma de Madrid (IIS-FJD, UAM), Madrid, Spain

^37^, Centre for Biomedical Network Research on Mental Health (CIBERSAM), Instituto de Salud Carlos III, Madrid, Spain

^38^, Fundación Hospital Infantil Universitario de San José, Bogotá, Colombia

^39^, Fundación Universitaria de Ciencias de la Salud, Bogotá, Colombia

^40^, Universidade Federal do Rio Grande do Norte, Programa de Pós-graduação em Ciências da Saúde, Natal, Brazil

^41^, Universidade Federal do Rio Grande do Norte, Departamento de Medicina Clínica, Natal, Brazil

^42^, Departamento de Genética e Morfologia, Instituto de Ciências Biológicas, Universidade de Brasília, Brasilia, Brazil

^43^, Colégio Marista de Brasilia, Brazil

^44^, Associação Brasileira de Educação e Cultura, Brazil

^45^, IDIVAL, Santander, Spain

^46^, Universidad de Cantabria, Santander, Spain

^47^, Hospital U M Valdecilla, Santander, Spain

^48^, Hospital Universitario La Paz-IDIPAZ, Servicio de Medicina Interna, Madrid, Spain

^49^, Fundació Docència I Recerca Mutua Terrassa, Barcelona, Spain

^50^, Spanish National Cancer Research Center, CNIO Biobank, Madrid, Spain

^51^, Fundación Santa Fe de Bogota, Departamento Patologia y Laboratorios, Bogotá, Colombia

^52^, Hospital General de Occidente, Zapopan, Jalisco, Mexico

^53^, Centro Universitario de Tonalá, Universidad de Guadalajara, Tonalá, Jalisco, Mexico

^54^, Centro de Investigación Multidisciplinario en Salud, Universidad de Guadalajara, Tonalá, Jalisco, Mexico

^55^, Instituto Murciano de Investigación Biosanitaria (IMIB-Arrixaca), Murcia, Spain

^56^, Universidad Católica San Antonio de Murcia (UCAM), Murcia, Spain

^57^, Hospital Universitario de Salamanca-IBSAL, Servicio de Medicina Interna-Unidad de Enfermedades Infecciosas, Salamanca, Spain

^58^, Universidad de Salamanca, Salamanca, Spain

^59^, Hospital Universitario de Fuenlabrada, Department of Internal Medicine, Madrid, Spain

^60^, Escola Tecnica de Saúde, Laboratorio de Vigilancia Molecular Aplicada, Pará, Brazil

^61^, Federal University of Pernambuco, Genetics Postgraduate Program, Recife, PE, Brazil

^62^, Hospital Universitario Infanta Leonor, Servicio de Alergia, Madrid, Spain

^63^, Hospital Universitario del Tajo, Servicio de Medicina Intensiva, Aranjuez, Spain

^64^, Hospital Universitario Mutua Terrassa, Barcelona, Spain

^65^, Hospital Universitario La Paz-IDIPAZ, Servicio de Farmacología, Madrid, Spain

^66^, Alcaldía de Barranquilla, Secretaría de Salud, Barranquilla, Colombia

^67^, Instituto de Investigación Sanitaria de Santiago (IDIS), Xenética Cardiovascular, Santiago de Compostela, Spain

^68^, Centre for Biomedical Network Research on Cardiovascular Diseases (CIBERCV), Instituto de Salud Carlos III, Madrid, Spain

^69^, Unidad de Infección Viral e Inmunidad, Centro Nacional de Microbiología (CNM), Instituto de Salud Carlos III (ISCIII), Madrid, Spain

^70^, Cardiovascular Genetics Center, Institut d’Investigació Biomèdica Girona (IDIBGI), Girona, Spain

^71^, Medical Science Department, School of Medicine, University of Girona, Girona, Spain

^72^, Hospital Josep Trueta, Cardiology Service, Girona, Spain

^73^, Institute of Biomedicine of Seville (IBiS), Consejo Superior de Investigaciones Científicas (CSIC)- University of Seville- Virgen del Rocio University Hospital, Seville, Spain

^74^, Division of Infectious Diseases, Instituto de Investigación Sanitaria-Fundación Jiménez Díaz University Hospital - Universidad Autónoma de Madrid (IIS-FJD, UAM), Madrid, Spain

^75^, Intensive Care Unit, Hospital Universitario Insular de Gran Canaria, Las Palmas de Gran Canaria, Spain

^76^, Hospital Universitario Mutua Terrassa, Terrassa, Spain

^77^, Departamento de Medicina, Hospital Universitario Virgen del Rocío,Universidad de Sevilla, Seville, Spain

^78^, Centre for Biomedical Network Research on Epidemiology and Public Health (CIBERESP), Instituto de Salud Carlos III, Madrid, Spain

^79^, Institute of Biomedicine of Seville (IBiS), Consejo Superior de Investigaciones Científicas (CSIC)- University of Seville- Virgen del Rocio University Hospital, Seville, Spain

^80^, Universidad de los Andes, Facultad de Ciencias, Bogotá, Colombia

^81^, Hospital Universitario de Salamanca-IBSAL, Servicio de Medicina Interna, Salamanca, Spain

^82^, Tecnologico de Monterrey, Escuela de Medicina y Ciencias de la Salud and Hospital San Jose TecSalud, Monterrey, Mexico

^83^, University of Fortaleza (UNIFOR), Department of Nutrition. Fortaleza, Brazil

^84^, Departamento de Química, Faculdade de Filosofia, Ciências e Letras de Ribeirão Preto, Universidade de São Paulo, Brazil

^85^, Andalusian Public Health System Biobank, Granada, Spain

^86^, Universidade Federal do Rio Grande do Norte, Programa de Pós-Graduação em Ciências Farmacêuticas, Natal, Brazil

^87^, Neuromuscular Unit, Neurology Department, Hospital Universitari de Bellvitge, L’Hospitalet de Llobregat (Barcelona), Spain

^88^, Bellvitge Biomedical Research Institute (IDIBELL), Neurometabolic Diseases Laboratory, L’Hospitalet de Llobregat, Spain

^89^, Centre for Biomedical Network Research on Diabetes and Metabolic Associated Diseases (CIBERDEM), Instituto de Salud Carlos III, Madrid, Spain

^90^, University of Pais Vasco, UPV/EHU, Bizkaia, Spain

^91^, Oncology and Genetics Unit, Instituto de Investigacion Sanitaria Galicia Sur, Xerencia de Xestion Integrada de Vigo-Servizo Galego de Saúde, Vigo, Spain

^92^, Hospital Universitario La Paz, Hospital Carlos III, Madrid, Spain

^93^, Hospital de San José, Sociedad de Cirugía de Bogota, Bogotá, Colombia

^94^, Hospital Universitario Río Hortega, Valladolid, Spain

^95^, Servicio de Medicina intensiva, Complejo Hospitalario Universitario de A Coruña (CHUAC), Sistema Galego de Saúde (SERGAS), A Coruña, Spain

^96^, Valencia University, Preventive Medicine Department, Valencia, Spain

^97^, Centre for Biomedical Network Research on Physiopatology of Obesity and Nutrition (CIBEROBN), Instituto de Salud Carlos III, Madrid, Spain

^98^, Research Unit, Hospital Universitario Ntra. Sra. de Candelaria, Instituto de Investigación Sanitaria de Canarias, Santa Cruz de Tenerife, Spain

^99^, Centre for Biomedical Network Research on Respiratory Diseases (CIBERES), Instituto de Salud Carlos III, Madrid, Spain

^100^, Otto von Guericke University, Departament of Microgravity and Translational Regenerative Medicine, Magdeburg, Germany

^101^, Maternidade Escola Janário Cicco, Natal, Brazil

^102^, Centro Singular de Investigación en Medicina Molecular y Enfermedades Crónicas (CIMUS), Universidade de Santiago de Compostela, Santiago de Compostela, Spain

^103^, Institute of Psychiatry and Mental Health, Hospital General Universitario Gregorio Marañón (IiSGM), Madrid, Spain

^104^, Programa de Pós Graduação em Ciências da Saúde, Faculdade de Medicina, Universidade de Brasília, Brasilia, Brazil

^105^, Fundació Docència I Recerca Mutua Terrassa, Terrassa, Spain

^106^, Hospital Universitario Mostoles, Unidad de Genética, Madrid, Spain

^107^, Instituto Aragonés de Ciencias de la Salud (IACS), Zaragoza, Spain

^108^, Instituto Investigación Sanitaria Aragón (IIS-Aragon), Zaragoza, Spain

^109^, Department of Preventive Medicine and Public Health, School of Medicine, Universidad Autónoma de Madrid, Madrid, Spain

^110^, IdiPaz (Instituto de Investigación Sanitaria Hospital Universitario La Paz), Madrid, Spain

^111^, Hospital Universitario Virgen del Rocío, Servicio de Medicina Interna, Seville, Spain

^112^, Unidad Diagnóstico Molecular. Fundación Rioja Salud, La Rioja, Spain

^113^, Hospital Universitario Quironsalud Madrid, Madrid, Spain

^114^, Hospital Universitario de Salamanca-IBSAL, Servicio de Cardiología, Salamanca, Spain

^115^, Hospital Universitario Puerta de Hierro, Servicio de Medicina Interna, Majadahonda, Spain

^116^, Biocruces Bizkaia Health Research Institute, Galdakao University Hospital, Osakidetza, Bizkaia, Spain

^117^, Instituto Regional de Investigación en Salud-Universidad Nacional de Caaguazú, Caaguazú, Paraguay

^118^, Universidade Federal do Pará, Núcleo de Pesquisas em Oncologia, Belém, Pará, Brazil

^119^, Hospital Ophir Loyola, Departamento de Ensino e Pesquisa, Belém, Pará, Brazil

^120^, Universidad Nacional de Asunción, Facultad de Politécnica, Paraguay

^121^, Fundación Asilo San Jose, Santander, Spain

^122^, Unidad de Enfermedades Infecciosas, Servicio de Medicina Interna, Hospital Universitario Puerta de Hierro, Instituto de Investigación Sanitaria Puerta de Hierro - Segovia de Arana, Madrid, Spain

^123^, Urgencias Hospitalarias, Complejo Hospitalario Universitario de A Coruña (CHUAC), Sistema Galego de Saúde (SERGAS), A Coruña, Spain

^124^, Grupo de Investigación en Interacciones Gen-Ambiente y Salud (GIIGAS) - Instituto de Biomedicina (IBIOMED), Universidad de León, León, Spain

^125^, Hospital Universitario Niño Jesús, Pediatrics Department, Madrid, Spain

^126^, Unitat de Malalties Infeccioses i Importades, Servei de Pediatría, Infectious and Imported Diseases, Pediatric Unit, Hospital Universitari Sant Joan de Deú, Barcelona, Spain

^127^, Microbiology Department, Instituto de Investigación Sanitaria-Fundación Jiménez Díaz University Hospital - Universidad Autónoma de Madrid (IIS-FJD, UAM), Madrid, Spain

^128^, Fundación Pública Galega de Medicina Xenómica, Sistema Galego de Saúde (SERGAS) Santiago de Compostela, Spain

^129^, Instituto de Investigación Sanitaria de Santiago (IDIS), Santiago de Compostela, Spain

^130^, Instituto de Genética Médica y Molecular (INGEMM), Hospital Universitario La Paz-IDIPAZ, Madrid, Spain

^131^, Hospital de Niños Ricardo Gutierrez, Buenos Aires, Argentina

^132^, Centre for Biomedical Network Research on Rare Diseases (CIBERER), Instituto de Salud Carlos III, Madrid, Spain Universidad Francisco de Vitoria, Madrid,Spain

^133^, Hospital Infanta Elena, Servicio de Medicina Intensiva, Valdemoro, Madrid, Spain

^134^, University of Salamanca, Biomedical Research Institute of Salamanca (IBSAL), Salamanca, Spain

^135^, Department of Immunology, IRYCIS, Hospital Universitario Ramón y Cajal, Madrid, Spain

^136^, Osakidetza, Cruces University Hospital, Bizkaia, Spain

^137^, Instituto Mexicano del Seguro Social, IMSS. Centro Médico Nacional La Raza. Hospital de Infectología. Mexico City, Mexico.

^138^, Hospital Universitario de Getafe, Servicio de Genética, Madrid, Spain

^139^, Ministerio de Salud Ciudad de Buenos Aires, Buenos Aires, Argentina

^140^, Hospital Clinico Universitario de Valladolid, Unidad de Apoyo a la Investigación, Valladolid, Spain

^141^, Universidad de Valladolid, Departamento de Cirugía, Valladolid, Spain

^142^, Secretaria Municipal de Saude de Apodi, Natal, Brazil

^143^, Sección Genética Médica - Servicio de Pediatría, Hospital Clínico Universitario Virgen de la Arrixaca, Servicio Murciano de Salud, Murcia, Spain

^144^, Departamento Cirugía, Pediatría, Obstetricia y Ginecología, Facultad de Medicina, Universidad de Murcia (UMU), Murcia, Spain

^145^, Hospital Universitario Centro Dermatológico Federico Lleras Acosta, Bogotá, Colombia

^146^, Hospital Universitario Virgen de las Nieves, Servicio de Análisis Clínicos e Inmunología, Granada, Spain

^147^, Pneumology Department, Hospital General Universitario Gregorio Marañón (iiSGM), Madrid, Spain

^148^, Intermediate Respiratory Care Unit, Department of Pneumology, Instituto de Investigación Sanitaria-Fundación Jiménez Díaz University Hospital - Universidad Autónoma de Madrid (IIS-FJD, UAM), Madrid, Spain

^148^, Clinica Comfamiliar Risaralda, Pereira, Colombia

^150^, Centro Universitario de Tonalá, Universidad de Guadalajara, Guadalajara, Mexico

^151^, Centro de Investigación Multidisciplinario en Salud, Universidad de Guadalajara, Guadalajara, Mexico

^152^, Unidad de Cuidados, Intensivos Hospital Clínico Universitario de Santiago (CHUS), Sistema Galego de Saúde (SERGAS), Santiago de Compostela, Spain

^153^, IIS La Fe, Plataforma de Farmacogenética, Valencia, Spain

^154^, Universidad de Valencia, Departamento de Farmacología, Valencia, Spain

^155^, Data Analysis Department, Instituto de Investigación Sanitaria-Fundación Jiménez Díaz University Hospital - Universidad Autónoma de Madrid (IIS-FJD, UAM), Madrid, Spain

^156^, Hospital del Mar, Infectious Diseases Service, Barcelona, Spain

^157^, Institut Hospital del Mar d’Investigacions Mèdiques (IMIM), Barcelona, Spain

^158^, CEXS-Universitat Pompeu Fabra, Spanish Network for Research in Infectious Diseases (REIPI), Barcelona, Spain

^159^, Biocruces Bizkaia Health Research Institute, Basurto University Hospital, Osakidetza, Bizkaia, Spain

^160^, Sabin Medicina Diagnóstica, Brazil

^161^, Opthalmology Department, Instituto de Investigación Sanitaria-Fundación Jiménez Díaz University Hospital - Universidad Autónoma de Madrid (IIS-FJD, UAM), Madrid, Spain

^162^, Hospital Sant Joan de Deu,Pediatric Critical Care Unit, Barcelona, Spain

^163^, Paediatric Intensive Care Unit, Agrupación Hospitalaria Clínic-Sant Joan de Déu, Esplugues de Llobregat, Barcelona, Spain

^164^, Hospital Universitario 12 de Octubre, Department of Immunology, Madrid, Spain

^165^, Instituto de Investigación Sanitaria Hospital 12 de Octubre (imas12), Transplant Immunology and Immunodeficiencies Group, Madrid, Spain

^166^, SIGEN Alianza Universidad de los Andes - Fundación Santa Fe de Bogotá, Bogotá, Colombia

^167^, Hospital General de Segovia, Medicina Intensiva, Segovia, Spain

^168^, Programa de Pós-Graduação em Biologia Animal, Universidade de Brasília, Brasília, Brazil

^169^, Clinical Trials Unit, Instituto de Investigación Sanitaria-Fundación Jiménez Díaz University Hospital - Universidad Autónoma de Madrid (IIS-FJD, UAM), Madrid, Spain

^170^, IMDEA-Food Institute, CEI UAM+CSIC, Madrid, Spain

^171^, Intensive Care Unit, Hospital Universitario de Canarias, La Laguna, Spain

^172^, Dirección General de Salud Pública, Consejería de Sanidad, Junta de Castilla y León, Valladolid, Spain

^173^, Universidade Federal do Rio Grande do Norte, Departamento de Analises Clinicas e Toxicologicas, Natal, Brazil

^174^, Hospital Universitario La Paz-IDIPAZ, Servicio de Inmunología, Madrid, Spain

^175^, La Paz Institute for Health Research (IdiPAZ), Lymphocyte Pathophysiology in Immunodeficiencies Group, Madrid, Spain

^176^, Hospital Universitario Virgen de las Nieves, Servicio de Enfermedades Infecciosas, Granada, Spain

^177^, Instituto de Investigación Biosanitaria de Granada (ibs GRANADA), Granada, Spain

^178^, Universidad de Granada, Departamento de Medicina, Granada, Spain

^179^, Faculdade de Medicina, Universidade de Brasília, Brasilia, Brazil

^180^, Fundación Jiménez Díaz, Epidemiology, Madrid, Spain

^181^, Universidad Autónoma de Madrid, Department of Medicine, Madrid, Spain

^182^, Universidad de Valladolid, Departamento de Medicina, Valladolid, Spain

^183^, Hospital Universitario Infanta Leonor, Servicio de Medicina Intensiva, Madrid, Spain

^184^, Unidad de Genética y Genómica Islas Baleares, Islas Baleares, Spain

^185^, Hospital Universitario Son Espases, Unidad de Diagnóstico Molecular y Genética Clínica, Islas Baleares, Spain

^186^, Genomics of Complex Diseases Unit, Research Institute of Hospital de la Santa Creu i Sant Pau, IIB Sant Pau, Barcelona, Spain

^187^, Intensive Care Unit, Hospital Universitario Ntra. Sra. de Candelaria, Santa Cruz de Tenerife, Spain

^188^, Preventive Medicine Department, Instituto de Investigación Sanitaria-Fundación Jiménez Díaz University Hospital - Universidad Autónoma de Madrid (IIS-FJD, UAM), Madrid, Spain

^189^, Servicio de Medicina Interna, Sanatorio Franchin, Buenos Aires, Argentina

^190^, Hospital Universitario del Tajo, Servicio de Medicina Intensiva, Toledo, Spain

^191^, Programa de Pós-Graduação em Ciências Médicas, Universidade de Brasília, Brasilia, Brazil

^192^, Programa de Pós-Graduação em Ciências da Saúde, Universidade de Brasília, Brasilia, Brazil

^193^, Hospital El Bierzo, Gerencia de Asistencia Sanitaria del Bierzo (GASBI), Gerencia Regional de Salud (SACYL), Ponferrada, Spain

^194^, Grupo INVESTEN, Instituto de Salud Carlos III, Madrid, Spain

^195^, Unidad de Cuidados Intensivos, Complejo Universitario de A Coruña (CHUAC), Sistema Galego de Saúde (SERGAS), A Coruña, Spain

^196^ Programa de Pós-Graduação em Genética da Faculdade de Medicina de Ribeirão Preto

^197^ Programa de Pós-Graduação em Química da Faculdade de Filosofia, Ciências e Letras de Ribeirão Preto

^198^, Hospital El Bierzo, Unidad Cuidados Intensivos, León, Spain

^199^, Spanish National Cancer Research Centre, Familial Cancer Clinical Unit, Madrid, Spain

^200^, Instituto de Investigación Sanitaria San Carlos (IdISSC), Hospital Clínico San Carlos (HCSC), Madrid, Spain

^201^, Hospital Universitario Severo Ochoa, Servicio de Medicina Interna, Madrid, Spain

^202^, Universidad de Sevilla, Departamento de Enfermería, Seville, Spain

^203^, Hospital General Universitario Gregorio Marañón (IiSGM), Madrid, Spain

^204^, Hospital Universitario La Paz-IDIPAZ, Servicio de Pediatría, Madrid, Spain

^205^, ERN-ITHACA-European Reference Network

^206^, Unidad de Genética y Genómica Islas Baleares, Unidad de Diagnóstico Molecular y Genética Clínica, Hospital Universitario Son Espases, Islas Baleares, Spain

^207^, Instituto de Investigación Sanitaria Islas Baleares (IdISBa), Islas Baleares, Spain

^208^, Programa de Pós-Graduação em Biologia Animal, Universidade de Brasília, Brasília, Brazil

^209^, Programa de Pós-Graduação em Ciências da Saúde, Universidade de Brasília, Brasília, Brazil

^210^, Programa de Pós-Graduação Profissional em Ensino de Biologia, Universidade de Brasília, Brasília, Brazil

^211^, Programa de Pós-Graduação em Ciências Médicas, Universidade de Brasília, Brasília, Brazil

^212^, Anatomía Patológica, Instituto de Investigación Sanitaria San Carlos (IdISSC), Hospital Clínico San Carlos (HCSC), Madrid, Spain

^213^, Tecnológico de Monterrey, Monterrey, Mexico

^214^, Infectious Diseases, Microbiota and Metabolism Unit, CSIC Associated Unit, Center for Biomedical Research of La Rioja (CIBIR), Logroño, Spain

^215^, Centro de Investigación en Anomalías Congénitas y Enfermedades Raras (CIACER), Universidad Icesi

^216^, Departamento de Genetica, Fundación Valle del Lili

^217^, Universidad Complutense de Madrid, Department of Immunology, Ophthalmology and ENT, Madrid, Spain

^218^, Department of Neumology, Instituto de Investigación Sanitaria-Fundación Jiménez Díaz University Hospital - Universidad Autónoma de Madrid (IIS-FJD, UAM), Madrid, Spain

^219^, Hospital Nuestra Señora de Sonsoles, Ávila, Spain

^220^, Inditex, A Coruña, Spain

^221^, Osakidetza, Cruces University Hospital, Barakaldo, Bizkaia, Spain

^222^, GENYCA, Madrid, Spain

^223^, Neuromuscular Diseases Unit, Department of Neurology, Hospital de la Santa Creu i Sant Pau, Universitat Autònoma de Barcelona, Barcelona, Spain

^224^, Instituto Mexicano del Seguro Social (IMSS), Centro Médico Nacional Siglo XXI, Unidad de Investigación Médica en Enfermedades Infecciosas y Parasitarias, Mexico City, Mexico

^225^, Catalan Institution of Research and Advanced Studies (ICREA), Barcelona, Spain

^226^, Intensive Care Department, Instituto de Investigación Sanitaria-Fundación Jiménez Díaz University Hospital - Universidad Autónoma de Madrid (IIS-FJD, UAM), Madrid, Spain

^227^, Hospital Universitario Príncipe de Asturias, Servicio de Microbiología Clínica, Madrid, Spain

^228^, Universidad de Alcalá de Henares, Departamento de Biomedicina y Biotecnología, Facultad de Medicina y Ciencias de la Salud, Madrid, Spain

^229^, Drug Research Centre, Institut d’Investigació Biomèdica Sant Pau, IIB-Sant Pau, Barcelona, Spain

^230^, Departamento de Genetica, Clinica imbanaco

^231^, Unidad de Cuidados Intensivos, Hospital Clínico Universitario de Santiago (CHUS), Sistema Galego de Saúde (SERGAS), Santiago de Compostela, Spain

^232^, Universidade de Brasília, Brasilia, Brazil

^233^, Department of Immunology, Hospital Universitario de Gran Canaria Dr. Negrín, Las Palmas de Gran Canaria, Spain

^234^, Department of Clinical Sciences, University Fernando Pessoa Canarias, Las Palmas de Gran Canaria, Spain

^235^, University Hospital Germans Trias i Pujol, Pediatrics Department, Badalona, Spain

^236^, Department of Pathology, Biobank, Instituto de Investigación Sanitaria-Fundación Jiménez Díaz University Hospital - Universidad Autónoma de Madrid (IIS-FJD, UAM), Madrid, Spain

^237^, Hospital Universitario Virgen de las Nieves, Servicio de Medicina Interna, Granada, Spain

^238^, Fundación Universitaria de Ciencias de la Salud, Grupo de Ciencias Básicas en Salud (CBS), Bogotá, Colombia

^239^, Sociedad de Cirugía de Bogotá, Hospital de San José, Bogotá, Colombia

^240^, Universidad de Granada, Departamento Bioquímica, Biología Molecular e Inmunología III, Granada, Spain

^241^, Hospital Infanta Elena, Allergy Unit, Valdemoro, Madrid, Spain

^242^, Faculty of Medicine, Universidad Francisco de Vitoria, Madrid, Spain

^243^, Hospital Universitario Infanta Leonor, Madrid, Spain

^244^, Complutense University of Madrid, Madrid, Spain

^245^, Gregorio Marañón Health Research Institute (IiSGM), Madrid, Spain

^246^ Colégio Militar de Brasília

^247^, The John Walton Muscular Dystrophy Research Centre, Newcastle University and Newcastle Hospitals NHS Foundation Trust, Newcastle upon Tyne, UK.

^248^, Neuromuscular Unit, Neuropediatrics Department, Institut de Recerca Sant Joan de Déu, Hospital Sant Joan de Déu, Spain

^249^, Fundació Docència i Recerca Mutua Terrassa, Terrassa, Spain

^250^, Casa de Saúde São Lucas, Natal, Brazil

^251^, Hospital Rio Grande, Rio Grande do Norte, Natal, Brazil

^252^, Universidade Federal do Rio Grande do Norte, Pós-graduação em Biotecnologia - Rede de Biotecnologia do Nordeste (Renorbio), Natal, Brazil

^253^, Intensive Care Unit, Hospital Universitario de Gran Canaria Dr. Negrín, Las Palmas de Gran Canaria, Spain

^254^, Universidad Fernando Pessoa Canarias, Las Palmas de Gran Canaria, Spain

^255^, Biobank, Puerta de Hierro-Segovia de Arana Health Research Institute, Madrid, Spain

^256^, Universidad Rey Juan Carlos, Madrid, Spain

^257^, Reumathology Service, Instituto de Investigación Sanitaria-Fundación Jiménez Díaz University Hospital - Universidad Autónoma de Madrid (IIS-FJD, UAM), Madrid, Spain

^258^, Hospital Clinico Universitario de Valladolid, Servicio de Anestesiologia y Reanimación, Valladolid, Spain

^259^, Hospital Clinico Universitario de Valladolid, Servicio de Hematologia y Hemoterapia, Valladolid, Spain

^260^, Hospital Universitario Lauro Wanderley, Brazil

^261^, Hospital Universitario Infanta Leonor, Servicio de Medicina Interna, Madrid, Spain

^262^, University Hospital of Burgos, Burgos, Spain

^263^, Universidad de Sevilla, Seville, Spain

^264^, Fundación Santa Fe de Bogota, Instituto de servicios medicos de Emergencia y trauma, Bogotá, Colombia

^265^, Universidad de los Andes, Bogotá, Colombia

^266^, Quironprevención, A Coruña, Spain

^267^, Junta de Castilla y León, Consejería de Sanidad, Valladolid, Spain

^268^, Gerencia Atención Primaria de Burgos, Burgos, Spain

^269^, Immunogenetics-Histocompatibility group, Servicio de Inmunología, Instituto de Investigación Sanitaria Puerta de Hierro - Segovia de Arana, Madrid, Spain

^270^, Hospital del Mar, Department of Infectious Diseases, Barcelona, Spain

^271^, IMIM (Hospital del Mar Medical Research Institute, Institut Hospital del Mar d’Investigacions Mediques), Barcelona, Spain

^272^, Universitat Autònoma de Barcelona, Department of Medicine, Spain

^273^, Consejería de Sanidad, Comunidad de Madrid, Madrid, Spain

^274^, Centro para el Desarrollo de la Investigación Científica, Asunción, Paraguay

^275^, Internal Medicine Department, Instituto de Investigación Sanitaria-Fundación Jiménez Díaz University Hospital - Universidad Autónoma de Madrid (IIS-FJD, UAM), Madrid, Spain

^276^, Universidade Federal do Rio Grande do Norte, Programa de Pós Graduação em Nutrição, Natal, Brazil

^277^, Preventive Medicine Department, Instituto de Investigacion Sanitaria Galicia Sur, Xerencia de Xestion Integrada de Vigo-Servizo Galego de Saúde, Vigo, Spain

^278^, Universidade Federal do Rio Grande do Norte, Departamento de Infectologia, Natal, Brazil

^279^, Hospital de Doenças Infecciosas Giselda Trigueiro, Rio Grande do Norte, Natal, Brazil

^280^, Genomics Division, Instituto Tecnológico y de Energías Renovables, Santa Cruz de Tenerife, Spain
